# Monocytic myeloid-derived suppressor cells, but not regulatory T cells, track immunoregulatory dynamics and relapse recovery in early RRMS

**DOI:** 10.64898/2026.05.25.26354018

**Authors:** Leticia Calahorra, Isabel Machín-Díaz, Inmaculada Alonso-García, José Manuel García-Domínguez, Inmaculada Pérez-Molina, Rafael Lebrón-Galán, Virginia Vila-del Sol, Haydee Goicoechea-Briceño, Jennifer García-Arocha, Rosa García-Montero, Victoria Galán, Guillermo Martín-Ávila, María Cabañas-Cotillas, María Cristina Ortega, Celia Camacho-Toledano, Mari Paz Serrano-Regal, Yolanda Aladro, María Luisa Martínez-Ginés, Diego Clemente

## Abstract

**Introduction:** Incomplete recovery from relapses contributes to long-term disability accumulation in relapsing–remitting multiple sclerosis (RRMS), yet the relationship between immune regulation and relapse recovery remains poorly defined.

**Objective:** To longitudinally characterize regulatory/effector immune cell dynamics in untreated RRMS patients and assess their association with immune balance and relapse recovery.

**Methods:** Monocytic myeloid-derived suppressor cells (M-MDSCs), regulatory T cells (Treg), and effector CD4⁺ T cell subsets were measured in blood from 69 untreated RRMS patients sampled during relapse or remission and re-evaluated after 12 months. Associations with clinical recovery after relapse were examined.

**Results:** During relapse, patients exhibited higher M-MDSC and Treg frequencies than in remission, while effector T cell subsets remained unchanged. Over one year, M-MDSCs increased consistently regardless of baseline clinical status, whereas Treg frequencies remained stable. Effector-to-M-MDSC ratios were markedly elevated during relapse and declined over time, while effector-to-Treg ratios showed minimal variation. M-MDSC levels during relapse were associated with sustained regulatory features at 12-month follow-up. Importantly, higher baseline M-MDSC levels, but not Treg frequencies, were associated with complete relapse recovery at one year.

**Conclusion:** These findings suggest that circulating M-MDSCs, but not Treg, reflect interindividual differences in immune regulation and clinical recovery after relapse in early RRMS.

## INTRODUCTION

Multiple sclerosis (MS) is a chronic disease in which the immune system attacks myelin and axons within the central nervous system (CNS), ultimately causing demyelination, axonal loss, and progressive disability[1]. Despite substantial advances in disease-modifying treatments (DMTs), relapse-associated worsening (RAW) remains a relevant contributor to disability accumulation in relapsing–remitting MS (RRMS) [2]. Longitudinal studies have shown that early disability accrual, including early RAW, may influence the subsequent risk of transition to secondary progressive MS (SPMS) [3]. However, the biological mechanisms underlying interindividual differences in relapse recovery remain poorly understood. Clinical predictors such as age and relapse severity only partially explain recovery outcomes [4], and reliable immunological correlates of favorable or unfavorable relapse recovery have not been clearly established. In this context, investigating peripheral immune dynamics during and after relapse may provide insight into mechanisms that support or limit neurological recovery.

In chronic diseases such as MS, alterations in circulating effector and regulatory immune cells can trigger aberrant immune responses, correlate with disease activity, and may serve as biomarkers[5]. CD4^+^ effector T helper (Th) cells, particularly Th1 and Th17, and the highly pro-inflammatory Th1.17 subset, contribute to MS pathophysiology, with Th17 up-regulated during relapses[6],[7]. Regulatory T cells (Treg) maintain immune homeostasis and limit effector T cell-mediated pathology [8]. However, their potential as predictors of relapse recovery in MS remains elusive.

In contrast to the well-defined role of T cells in MS pathophysiology, far less is known about the contribution of innate immunity, especially about those cells involved in immunoregulatory mechanisms. Myeloid-derived suppressor cells (MDSCs) is a heterogeneous population of immature myeloid cells that regulate immune responses via effector T cell suppression and/or Treg induction[9],[10]. Monocytic (M)-MDSCs have been associated with milder disease in MS, with circulating M-MDSCs correlating with improved relapse recovery in early, untreated RRMS [11], yet their interactions with effector T cells and Treg during the early phases of MS remain unexplored.

In this study, we investigated longitudinal peripheral immune dynamics in untreated RRMS patients sampled during relapse or remission. We focused on regulatory immune populations, including M-MDSCs and Treg subsets, to explore whether early immunoregulatory signatures are associated with subsequent clinical recovery.

## METHODS

### MS patient’s cohort

All participants had a prior MS according to the 2017 McDonald criteria[12]. The cohort comprised untreated RRMS patients recruited between 2015 and 2023 who had experienced their first relapse (MS debut) within the year prior to blood collection and had not received corticosteroids in the preceding 6 months (**Table 1**). Blood samples were obtained either during relapse or remission, in accordance with previously established criteria [13]. A relapse, qualified by a trained neurologist, was defined as a clinically meaningful episode of new neurological dysfunction lasting ≥ 24 hours, not attributable to fever or other illness, and separated from prior activity by at least 30 days, reflecting acute inflammatory demyelination. Patients were recruited from the University Hospital of Toledo (Toledo, Spain), Gregorio Marañón General University Hospital (Madrid, Spain) and Getafe University Hospital (Getafe, Spain). All study protocols were approved by the institutional ethics review boards, and written informed consent was obtained from all participants.

**Table 1.** Clinical/Demographic characteristics of MS patients.

|  | **RRMS**<br>**(n = 69)** | **Relapse**<br>**(n = 45)** | **Remission**<br>**(n = 24)** |
| --- | --- | --- | --- |
| Female (%)<sup>-</sup> | 48 (69.57) | 30 (65.23) | 17 (70.83) |
| Age (year)<sup>‡</sup> | 37 (30-44) | 36 (30-43) | 40 (34-45) |
| Baseline EDSS<sup>//</sup> | 3 (1-3) | 3 (2-3.5) * | 2 (1-3) * |
| 1 year follow-up EDSS<sup>//</sup> | 1 (1-1.5) | 1 (0-1) | 1 (0-2) |
| Baseline No. of brain MRI Gd+<sup>‡</sup> | 1.88 ± 0.14 | 2.12 ± 0.40 | 1.25 ± 0.47 |
| Baseline No brain MRI lesions on T2 (%)<sup>&</sup> |  |  |  |
| 0-9 lesions | 20 (28.99) | 10 (22.22) | 10 (41.47) |
| ≥ 10 lesions | 49 (71.01) | 35 (77.78) | 14 (58.33) |
| DMTs used during follow-up<sup>&</sup> |  |  |  |
| No treatment | 16 (23.19) | 11 (24.44) | 5 (20.83) |
| Injectable DMTs<sup>1</sup> | 10 (14.49) | 8 (16.33) | 2 (8.33) |
| Oral DMTs<sup>2</sup> | 39 (56.52) | 22 (44.90) | 17 (70.83) |
| Monoclonal antibodies<sup>3</sup> | 4 (5.80) | 4 (8.16) | 0 (0.00) |

<sup>‡</sup>The values are the mean ± SEM of each group.
<sup>//</sup>The values are the median (IQR).
\* Significant differences between groups. A p-value <0.05 was considered statistically significant. The p-values were calculated using the Mann-Whitney t-test for differences between groups (Relapse-Remission).
<sup>-</sup>The value is the number (percentage) of women in each group, and a $\chi^2$ exact test was used to compare two proportions.
<sup>&</sup>The value is the number (percentage) of patients in each group, and a Fisher's exact test (lesions) or Fisher-Freeman-Halton exact test (treatments) was used to compare the proportions.
<sup>1</sup> Injectable DMTs: glatiramer acetate, all interferon-β formulations.
<sup>2</sup> Oral DMTs: fumarates, teriflunomide, fingolimod, cladribine.
<sup>3</sup> Monoclonal antibodies: natalizumab, alemtuzumab.

### Sample collection

Peripheral blood mononuclear cells (PBMCs) were isolated from 15 mL blood using Ficoll density gradient centrifugation (GE-171440-02, Merck) at baseline and after 12-month follow-up. Cells were collected from the interphase, washed with isolation buffer (2.23 g/L D-glucose, 2.2 g/L sodium citrate, 0.8 g/L citric acid, 0.5% BSA in PBS) and centrifuged at 500 g for 10 minutes at room temperature. The resulting cell pellet was resuspended in fetal bovine serum (FBS), counted, and stored in a 1:1 mixture of FBS containing 20% DMSO (Sigma-Aldrich) at −165°C until their analysis.

### Flow Cytometry and Cell Staining

Cryopreserved PBMCs were thawed, counted, and 1 × 10^6^ cells per tube were stained for viability using Zombie-NIR dye (BioLegend), blocked with human IgG (Beriglobin) to prevent nonspecific Fc receptor binding and incubated with fluorochrome-conjugated antibodies. For intracellular staining, cells were fixed and permeabilized (BD Cytofix/Cytoperm™) before labeling with FoxP3. Stained samples were fixed in 2% paraformaldehyde (0.1.% for M-MDSCs) and stored at 4°C until acquisition on the same day (24 h later for M-MDSCs). Data were acquired using a FACS CANTO II cytometer and analyzed with FlowJo v10.7 (BD Life Sciences).

### Immune cell profiling

M-MDSCs were defined as CD14^+^CD15^−^CD11b^+^CD33^+^HLA-DR^−/low^[11]. Treg subsets were defined as CD4^+^CD25^hi^CD127^lo^FoxP3^+^, with CD39^+^ indicating stronger immunoregulatory capacity and CCR6^+^ indicating migratory potential to chronic inflammation sites [14],[15]. Effector CD4^+^ T cell subsets (Th1, Th17, Th1.17) were identified using CXCR3, CCR6, and CD161 markers, with CCR2 and CCR5 used to define T effector cells with high CNS-penetrating potential (Teff), and TeffCD161^+^ as Teff resistant to immunosuppression [16],[17]. Specific cell profiling, flow cytometry antibodies, and gating strategy can be found in **Supplementary Tables 1-2** and **Supplementary Figures 1-3**.

### Statistics

Data are presented as the mean ± SEM, or the median (interquartile range-IQR). Normality was assessed using the Shapiro–Wilk test. Comparisons between groups were carried out using one-way ANOVA or ANOVA on ranks, followed by Tukey or Dunn *post hoc* tests as appropriate. Comparisons between two groups were carried out using Student’s *t*-test or Mann–Whitney U test for non-parametric data. Paired *t*-test was used for comparison of cell subsets from baseline and follow-up sampling. Xi^2^ exact test was used to compare sex proportions. Fisher’s exact test or Fisher–Freeman–Halton exact test was used to compare the proportions of number of T2 lesins and treatments groups, respectively. Correlations were assessed using Pearson or Spearman tests as appropriate. A *p* < 0.05 was considered statistically significant (\**p* < 0.05; \*\**p* < 0.01; \*\*\**p* < 0.001).

## RESULTS

### Clinical Characteristics of RRMS patients

Sixty-nine untreated RRMS patients were included in the analysis. Baseline demographics and clinical characteristics are summarized in **Table 1**. Patients sampled during relapse showed higher baseline EDSS scores compared with those sampled in remission (p = 0.019), whereas no differences were observed in age (p = 0.233), sex distribution (p = 0.833), number of Gadolinium-enhancing lesions (p = 0.071), or number of T2 lesion load (p = 0.163). DMTs initiated during follow-up were similarly distributed between relapse-and remission-sampled patients (p = 0.247).

### Regulatory immune cells dynamically change between relapse and remission in untreated RRMS

We first examined longitudinal variations in immune phenotypes in untreated RRMS patients after their first relapse **(Table 2)**. Among CD4^+^ T cell subsets, only Th17 cells showed a negative correlation with days elapsed from relapse to sampling (r=-0.259; p=0.032). Regarding regulatory populations, M-MDSCs (r=-0.286; p=0.017), Treg (r=-0.241; p=0.047), and Treg CCR6^+^ (r=-0.283; p=0.019) percentages displayed similar behavior (**Figure 1A-D**). Comparing patients at relapse versus remission, no significant differences were observed in effector T cell subsets (**Figure 2A-B**). In contrast, regulatory cells, including M-MDSCs, total Treg and TregCCR6^+^ were significantly elevated during relapse compared with remission (**Figure 2C-E**). At 12-month follow-up, these differences were no longer present, and other T cell subsets remained unchanged.

**Figure 1.**
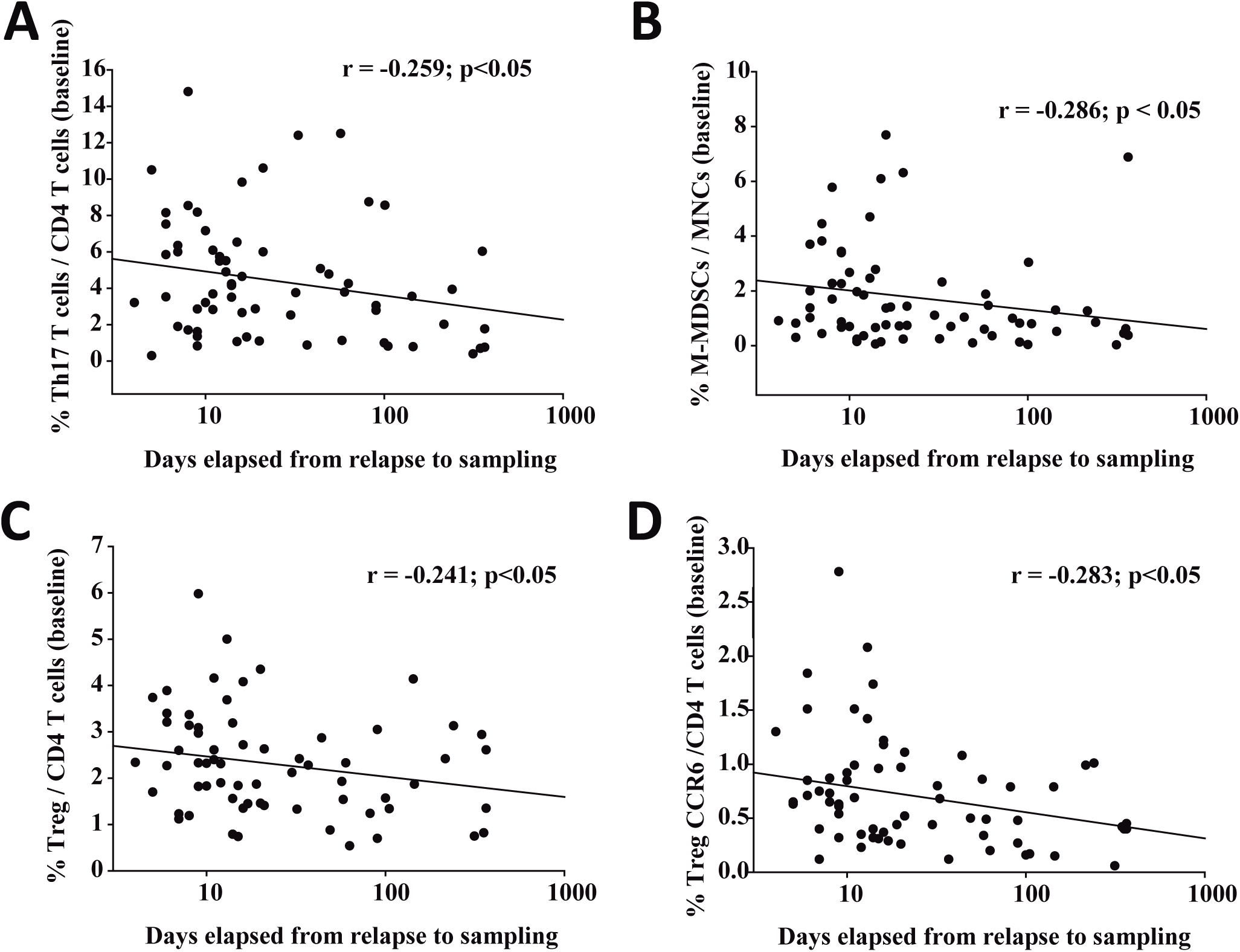
Correlation of immune cell frequencies at baseline with days elapsed since relapse. Frequencies of Th17/CD4⁺ T cells (A), M-MDSCs/mononuclear cells (B), total Treg/CD4⁺ T cells (C), and TregCCR6⁺/CD4⁺ T cells (D) were inversely correlated with time since relapse, indicating higher proportions closer to the inflammatory event.

**Figure 2.**
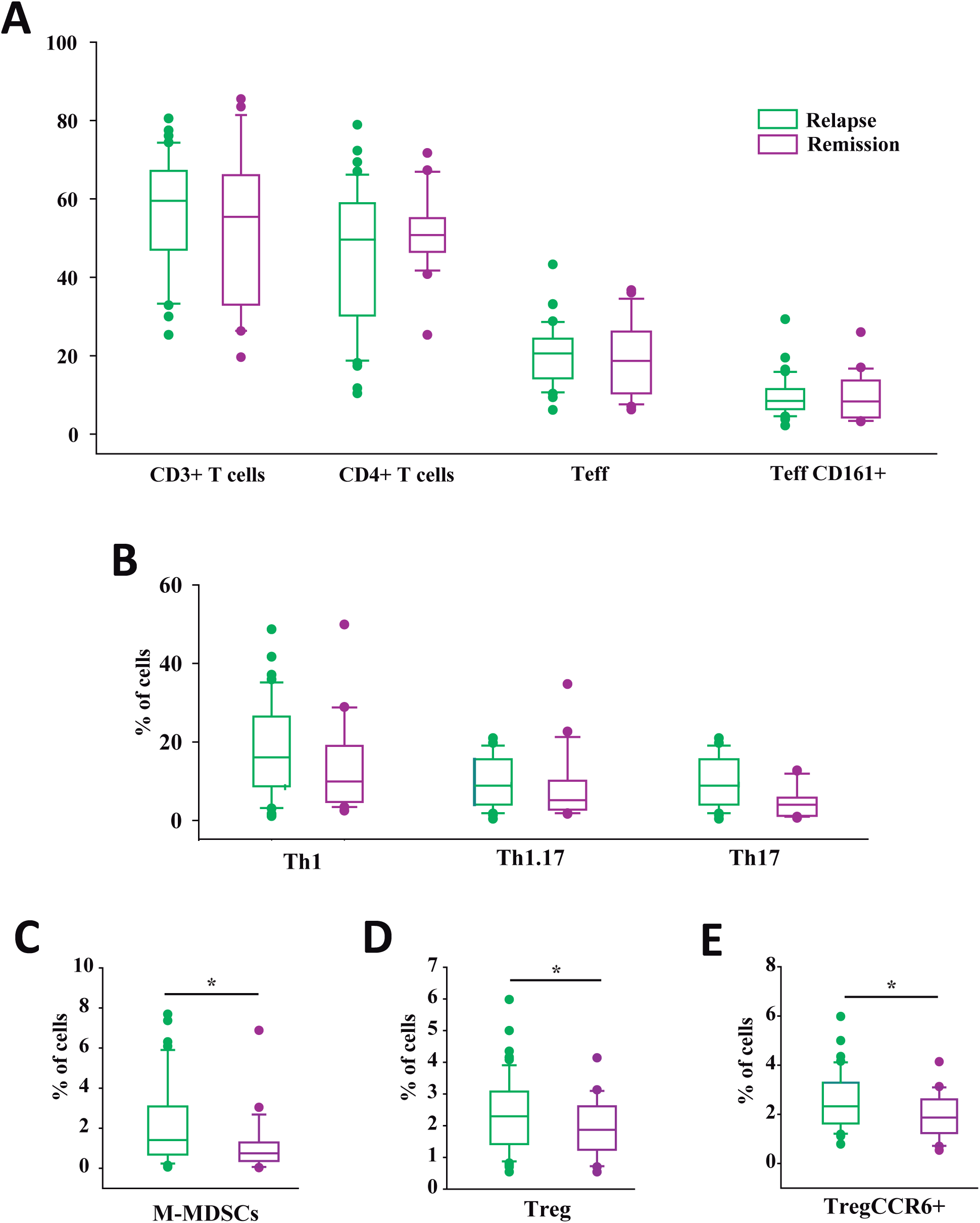
Comparison of immune cell frequencies between patients sampled during relapse or remission. No significant differences were detected in effector T cell subsets (A–B). In contrast, M-MDSCs among mononuclear cells (C), total Treg (D), and TregCCR6⁺ cells among CD4^+^ T cells (E) were significantly higher in patients sampled during relapse.

**Table 2.** Distribution of cell frequencies of MS untreated patients.

|  |  | **Relapse**<br>**(n = 45)** | **Remission**<br>**(n = 24)** | **p** |
| --- | --- | --- | --- | --- |
| **BASELINE** | M-MDSCs/MNCs | 1.41 (0.69-2.93) | 0.75 (0.37-1.29) | **0.016** |
|  | CD3<sup>+</sup>T cells | 59.50 (47.43-67.03) | 55.11 (44.50-66.20) | 0.427 |
|  | CD4<sup>+</sup>T cells | 49.60 (30.93-58.40) | 50.05 (45.98-54.98) | 0.348 |
|  | Teff | 8.30 (4.59-11.90) | 9.15 (5.72-12.30) | 0.469 |
|  | TeffCD161<sup>+</sup> | 3.58 (2.14-5.60) | 4.78 (2.58-6.36) | 0.239 |
|  | Th1 | 16.10 (8.98-26.13) | 10.85 (5.30-21.95) | 0.323 |
|  | Th1.17 | 8.62 (3.83-14.93) | 4.91 (2.52-9.69) | 0.104 |
|  | Th17 | 4.18 (2.42-6.40) | 3.31 (0.94-4.93) | 0.135 |
|  | Treg | 2.33 (1.67-3.25) | 1.87 (1.26-2.56) | **0.045** |
|  | TregCCR6<sup>+</sup> | 0.69 (0.40-1.02) | 0.45 (0.22-0.79) | **0.022** |
|  | TregCD39<sup>+</sup> | 0.96 (0.59-1.70) | 0.93 (0.45-1.57) | 0.513 |
| **ONE YEAR**<br>**FOLLOW-UP** | M-MDSCs/MNCs | 2.03 (1.23-4.92)*** | 2.05 (0.76-3.98)** | 0.461 |
|  | CD3<sup>+</sup>T cells | 53.20 (44.58-58.40)** | 47.30 (37.35-59.05) | 0.267 |
|  | CD4<sup>+</sup>T cells | 40.20 (30.70-53.88) | 52.45 (41.38-63.50) | 0.117 |
|  | Teff | 7.30 (4.05-9.14)* | 9.15 (5.72-12.30) | 0.724 |
|  | TeffCD161<sup>+</sup> | 2.90 (1.94-4.07)* | 3.41 (2.29-4.50) | 0.288 |
|  | Th1 | 15.40 (9.16-21.53) | 13.90 (7.55-19.90) | 0.865 |
|  | Th1.17 | 5.75 (3.06-25.15) | 4.60 (2.08-8.53) | 0.341 |
|  | Th17 | 3.30 (1.76-5.29)* | 2.44 (1.61-3.54) | 0.412 |
|  | Treg | 2.46 (1.85-3.38) | 2.25 (1.34-2.95) | 0.216 |
|  | TregCCR6<sup>+</sup> | 0.62 (0.42-0.92) | 0.52 (0.24-0.92) | 0.371 |
|  | TregCD39<sup>+</sup> | 1.13 (0.60-1.76) | 1.40 (0.50-2.06) | 0.881 |

\*The values are the median (IQR) of each group. \*p<0.05, \*\*p<0.01, \*\*\*p<0.001 vs. baseline.

Longitudinal analysis of effector T cells in patients sampled during relapse revealed a decreased over time in CD3⁺ T cells (p = 0.021), total Teff (p = 0.014), Teff CD161⁺ (p = 0.011), and Th17 cells (p = 0.042). Conversely, CD4^+^ T cells (p = 0.375), Th1 (p = 0.357), and Th1.17 (p = 0.099) frequencies remained stable (**Figure 3A-F**). Interestingly, among regulatory populations, only M-MDSCs increased over the 12-month follow-up (p < 0.001), whereas total Treg, TregCCR6^+^, and TregCD39^+^ did not show significant temporal changes (**Figure 3G-J**; **Table 2**). At one-year follow-up, most immune subsets were comparable between treated and untreated patients sampled during relapse, with differences observed only for Teff and total Treg frequencies (**Supplementary Table 3)**. Importantly, no treatment-by-time interactions were detected for any subset, arguing against treatment exposure as the main driver of the longitudinal changes observed. In patients sampled during remission, only M-MDSCs increased over one year (p = 0.002), while all other effector and regulatory populations remained stable (**Table 2**).

**Figure 3.**
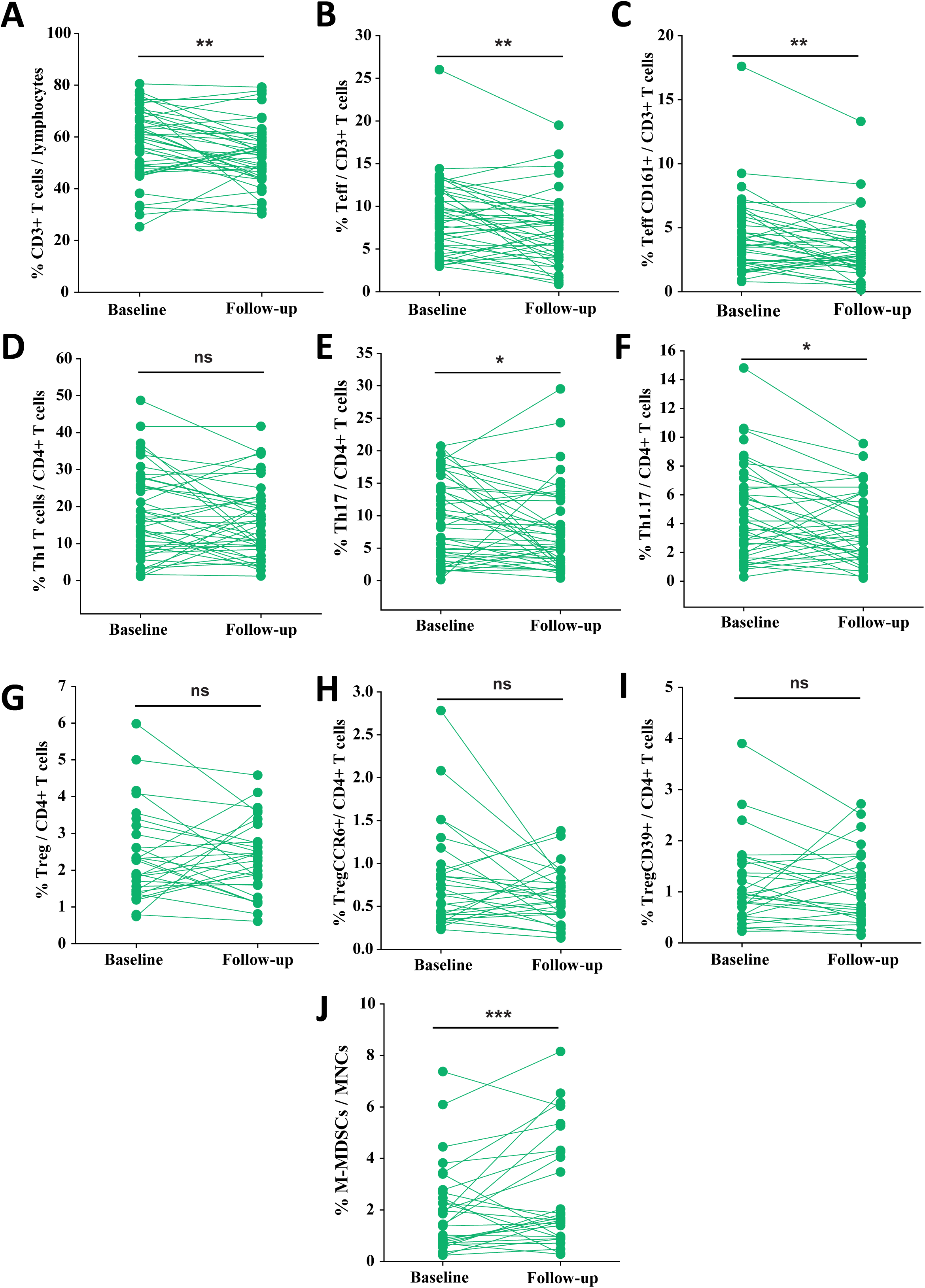
Longitudinal analysis of immune cell populations in patients sampled during relapse. CD3⁺ T cells (A), total Teff and TeffCD161⁺ cells (B–C), and Th17 and Th1.17 cells (E–F) decreased over 12 months. Th1 cells (D) and all analyzed Treg subsets (G–I) remained stable. Notably, M-MDSCs increased significantly over time (J).

These findings indicate that regulatory cells are the primary immune populations undergoing changes between relapse and remission, with M-MDSCs exhibiting consistent longitudinal dynamics, highlighting their potential role in shaping disease course in early RRMS.

To investigate the balance between regulatory populations and their effector T cell targets during relapse, we analyzed ratios of effector T cells to M-MDSCs and Tregs in untreated RRMS patients at baseline and 12-month follow-up. These ratios provide an estimate of the immunoregulatory environment, as effector T cells are the primary targets of M-MDSCs and Treg [18]. At baseline, patients sampled during relapse exhibited higher ratios of CD3⁺, CD4^+^, total Teff, and TeffCD161⁺ T cells relative to M-MDSCs compared with patients in remission (**Table 3**). One year later, these differences were no longer observed, and the vast majority of effector/M-MDSC ratios decreased compared with baseline (**Table 3**). In contrast, effector T cell/Treg ratios were largely stable over time. Differences between patients in relapse or remission were observed only in CD4^+^ /Treg at baseline (**Table 4**). Similar patterns were observed when using TregCCR6⁺ or TregCD39⁺ as reference populations (**Supplementary Tables 4 and 5**).

**Table 3.**
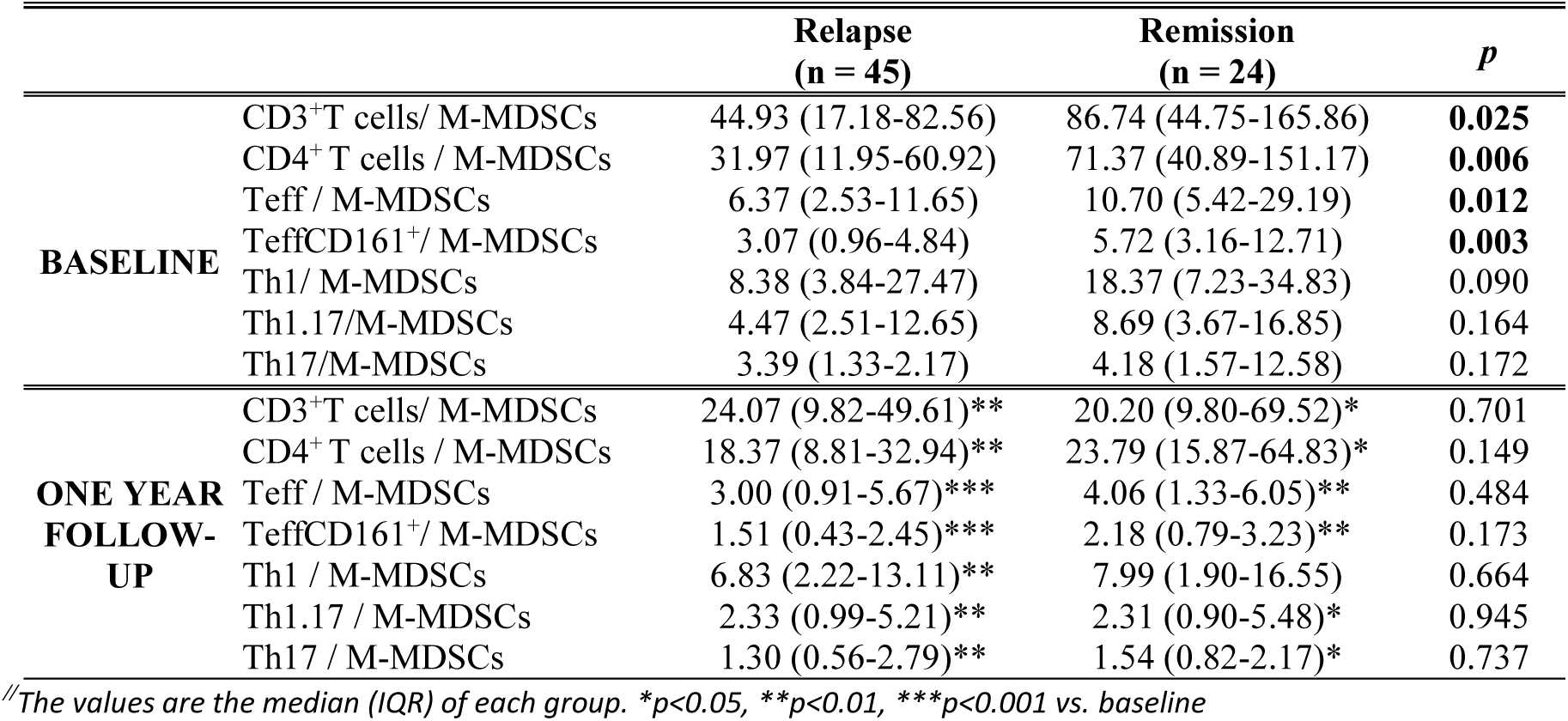
Distribution of effector T cells/M-MDSCs ratios in MS patients*^//^*.

|  |  | Relapse<br>(n = 45) | Remission<br>(n = 24) | *p* |
| --- | --- | --- | --- | --- |
| **BASELINE** | CD3<sup>+</sup>T cells/ M-MDSCs | 44.93 (17.18-82.56) | 86.74 (44.75-165.86) | **0.025** |
|  | CD4<sup>+</sup> T cells / M-MDSCs | 31.97 (11.95-60.92) | 71.37 (40.89-151.17) | **0.006** |
|  | Teff / M-MDSCs | 6.37 (2.53-11.65) | 10.70 (5.42-29.19) | **0.012** |
|  | TeffCD161<sup>+</sup>/ M-MDSCs | 3.07 (0.96-4.84) | 5.72 (3.16-12.71) | **0.003** |
|  | Th1/ M-MDSCs | 8.38 (3.84-27.47) | 18.37 (7.23-34.83) | 0.090 |
|  | Th1.17/M-MDSCs | 4.47 (2.51-12.65) | 8.69 (3.67-16.85) | 0.164 |
|  | Th17/M-MDSCs | 3.39 (1.33-2.17) | 4.18 (1.57-12.58) | 0.172 |
| **ONE YEAR**<br>**FOLLOW-**<br>**UP** | CD3<sup>+</sup>T cells/ M-MDSCs | 24.07 (9.82-49.61)** | 20.20 (9.80-69.52)* | 0.701 |
|  | CD4<sup>+</sup> T cells / M-MDSCs | 18.37 (8.81-32.94)** | 23.79 (15.87-64.83)* | 0.149 |
|  | Teff / M-MDSCs | 3.00 (0.91-5.67)*** | 4.06 (1.33-6.05)** | 0.484 |
|  | TeffCD161<sup>+</sup>/ M-MDSCs | 1.51 (0.43-2.45)*** | 2.18 (0.79-3.23)** | 0.173 |
|  | Th1 / M-MDSCs | 6.83 (2.22-13.11)** | 7.99 (1.90-16.55) | 0.664 |
|  | Th1.17 / M-MDSCs | 2.33 (0.99-5.21)** | 2.31 (0.90-5.48)* | 0.945 |
|  | Th17 / M-MDSCs | 1.30 (0.56-2.79)** | 1.54 (0.82-2.17)* | 0.737 |

<sup>//</sup>The values are the median (IQR) of each group. \**p*<0.05, \*\**p*<0.01, \*\*\**p*<0.001 vs. baseline

**Table 4.** Distribution of effector T cells/Treg ratios in MS patients*^//^*.

|  |  | **Relapse**<br>**(n = 45)** | **Remission**<br>**(n = 24)** | **p** |
| --- | --- | --- | --- | --- |
| **BASELINE** | CD3<sup>+</sup>T cells / Treg | 23.95 (14.81-34.41) | 28.31 (16.26-39.30) | 0.412 |
|  | CD4<sup>+</sup> T cells / Treg | 18.67 (12.72-27.12) | 23.44 (17.51-45.67) | **0.017** |
|  | Teff / Treg | 7.14 (4.04-12.49) | 10.31 (5.97-13.83) | 0.382 |
|  | TeffCD161<sup>+</sup>/ Treg | 2.62 (1.59-6.03) | 5.08 (3.45-7.82) | 0.066 |
|  | Th1/ Treg | 6.44 (3.55-11.72) | 7.02 (3.26-15.00) | 0.645 |
|  | Th1.17/ Treg | 3.79 (1.51-5.74) | 2.90 (1.12-4.69) | 0.552 |
|  | Th17/ Treg | 1.75 (1.01-3.19) | 0.89 (0.61-1.12) | 0.382 |
| **ONE YEAR**<br>**FOLLOW-UP** | CD3<sup>+</sup>T cells / Treg | 20.73 (15.43-29.13) | 23.29 (14.88-32.68) | 0.467 |
|  | CD4<sup>+</sup> T cells / Treg | 16.78 (11.69-26.76) | 21.32 (15.64-37.66) | 0.052 |
|  | Teff/ Treg | 6.54 (4.92-11.33) | 7.01 (4.62-12.67) | 0.580 |
|  | TeffCD161<sup>+</sup>/ Treg | 3.31 (2.30-4.20) | 3.22 (2.08-6.23) | 0.307 |
|  | Th1 / Treg | 6.13 (3.08-11.70) | 8.79 (3.89-9.81) | 0.227 |
|  | Th1.17 / Treg | 2.96 (1.28-4.94) | 2.11 (0.81-4.25) | 0.443 |
|  | Th17 / Treg | 1.34 (0.66-2.22)* | 1.01 (0.76-1.93) | 0.730 |

<sup>//</sup>The values are the median (IQR) of each group. \*p<0.05 vs. baseline

**Table 5.** Baseline clinical characteristics of relapsing MS patients with full or partial recovery.

|  | Relapse Partial<br>Recovery<br>(n = 23) | Relapse Full<br>Recovery<br>(n = 21) |
| --- | --- | --- |
| Female (%) | 13 (56.62) | 14 (66.67) |
| Age (year)<sup>‡</sup> | 39 (31-42) | 32 (24-44) |
| Baseline EDSS<sup>//</sup> | 3 (2-4)* | 3 (2-3)* |
| 1 year follow-up EDSS<sup>//</sup> | 1 (1-2) | 0 |
| Baseline No. of brain MRI Gd<sup>+</sup> lesions<sup>‡</sup> | 1.78 ± 0.50 | 2.85 ± 0.69 |
| Baseline No. of brain MRI lesions on T2<sup>&</sup> |  |  |
| 0-9 lesions | 5 (21.74) | 6 (28.57) |
| ≥ 10 lesions | 18 (78.26) | 15 (71.43) |
| DMTs used during follow-up<sup>&</sup> |  |  |
| No treatment | 5 (21.74) | 5 (23.81) |
| Injectable DMTs<sup>1</sup> | 5 (21.74) | 3 (14.29) |
| Oral DMTs<sup>2</sup> | 10 (43.48) | 12 (57.14) |
| Monoclonal antibodies<sup>3</sup> | 3 (13.04) | 1 (4.76) |

<sup>‡</sup>The values are the mean ± SEM of each group.
<sup>//</sup>The values are the median (IQR).
\* Significant differences between groups. The *p*-values were calculated using the Mann-Whitney *t*-test for differences between groups.
<sup>-</sup>The value is the percentage of women in each group, and a $\chi^2$ exact test was used to compare two proportions.
<sup>&</sup>The value is the number (percentage) of patients in each group, and a Fisher's exact test (lesions) or Fisher-Freeman-Halton exact test (treatments) was used to compare the proportions.
<sup>1</sup> Injectable DMTs: glatiramer acetate, all interferon-β formulations.
<sup>2</sup> Oral DMTs: fumarates, teriflunomide, fingolimod, cladribine.
<sup>3</sup> Monoclonal antibodies: natalizumab, alemtuzumab.

Overall, these findings reveal a more pro-inflammatory effector/regulatory environment during relapse than during remission and suggest that M-MDSCs offer a more sensitive readout of regulatory activity than Treg, underscoring their role in shaping immune homeostasis and relapse recovery in untreated RRMS patients.

### Baseline M-MDSC levels during relapse are associated with long-term immune regulatory changes

To determine whether baseline levels of regulatory cells are associated with the effector and regulatory immune landscape over time, we evaluated correlations between baseline M-MDSC and Treg abundance and immune cell subsets measured at the same time point and after 12-month follow-up. At baseline, untreated RRMS patients sampled during relapse showed a positive correlation between M-MDSC levels and pro-inflammatory TeffCD161^+^ (r = 0.336, p = 0.024) and Th17 cells (r = 0.423, p = 0.004), with no significant correlation observed with other effector populations. In contrast, no correlation was detected between M-MDSCs and effector T cell types in patients sampled during remission. Interestingly, higher baseline M-MDSCs frequencies in patients sampled during relapse were associated with lower total CD4^+^ T cell proportions (r = -0.303, p = 0.042) and sustained enrichment of Th17 cells (r = 0.448, p = 0.004) at 12 months (**Figure 4A-B**). These associations were not observed for other effector subsets and were absent in patients sampled during remission, indicating a relapse-specific imprinting of immune trajectories by M-MDSCs. In contrast to M-MDSCs, baseline total Treg frequencies did not correlate with effector T cell subsets at baseline and were only inversely associated with Th17 cell frequencies at 12 months in patients sampled during relapse (r = -0.337, p = 0.033).

**Figure 4.**
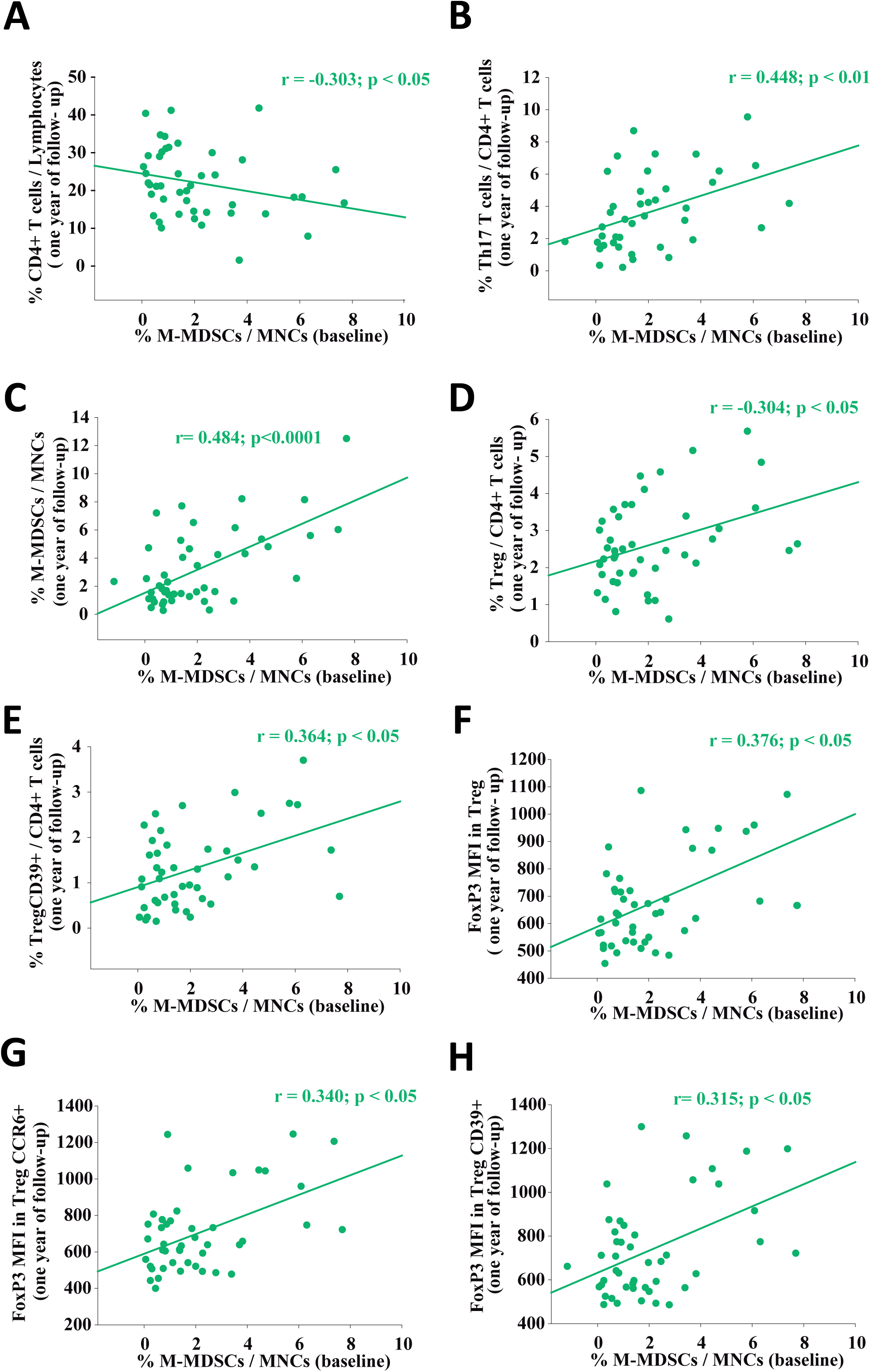
Associations between baseline M-MDSC levels and immune cell frequencies at 12-month follow-up. Baseline M-MDSCs correlated negatively with CD4⁺ T cells (A) and positively with Th17 cells (B). Higher baseline M-MDSC levels were associated with increased frequencies of M-MDSCs (C), total Treg (D), and TregCD39⁺ cells (E), as well as enhanced FoxP3 expression in total Treg, TregCCR6⁺, and TregCD39⁺ subsets (F–G).

Notably, in the same patients, baseline M-MDSC levels positively correlated with their frequencies at follow-up (r = 0.484, p = 0.00083), suggesting maintenance of this population over time (**Figure 4C**). In addition, baseline M-MDSC abundance was associated with higher frequencies of total Treg (r = 0.324, p = 0.043; **Figure 4D**) and with highly immunosuppressive TregCD39^+^ (r = 0.364, p = 0.014; **Figure 4E**), as well as increased FoxP3 expression (a marker associated with Treg activity in MS patients; [17]) within Treg populations at follow-up (total Treg: r = 0.376, p = 0.011; TregCCR6^+^: r = 0.340, p = 0.023; TregCD39^+^: r = 0.315, p = 0.035; **Figure 4F-H**). These associations were not observed in patients sampled during remission.

Overall, these findings show that baseline circulating M-MDSC levels, rather than Treg, were associated with regulatory immune features detected 12 months later in untreated RRMS patients sampled during relapse.

### Baseline M-MDSCs, but not Treg, are associated with favorable relapse recovery in RRMS

We aimed to determine whether baseline regulatory cell levels differed according to recovery outcome compared to patients in remission. Relapse patients experienced either full recovery (EDSS = 0, n = 23) or partial recovery (EDSS > 0, n = 21) after 12-month follow-up. Baseline clinical characteristics were similar between the two groups, except a higher EDSS in patients with partial recovery (**Table 5**).

No differences were observed in effector T cell subsets among those patients and patients sampled during remission (**Table 6**). In contrast, M-MDSC levels were higher in patients who subsequently achieved full recovery compared to those in remission, whereas patients with partial recovery did not differ from those in remission (**Figure 5A**). Similarly, ratios of effector T cells to M-MDSCs were lower in patients who subsequently experienced full recovery (**Figure 5B-E**). In contrast, Treg frequencies and effector T cell/Treg ratios were comparable across groups (**Figure 5F–J)**.

**Figure 5.**
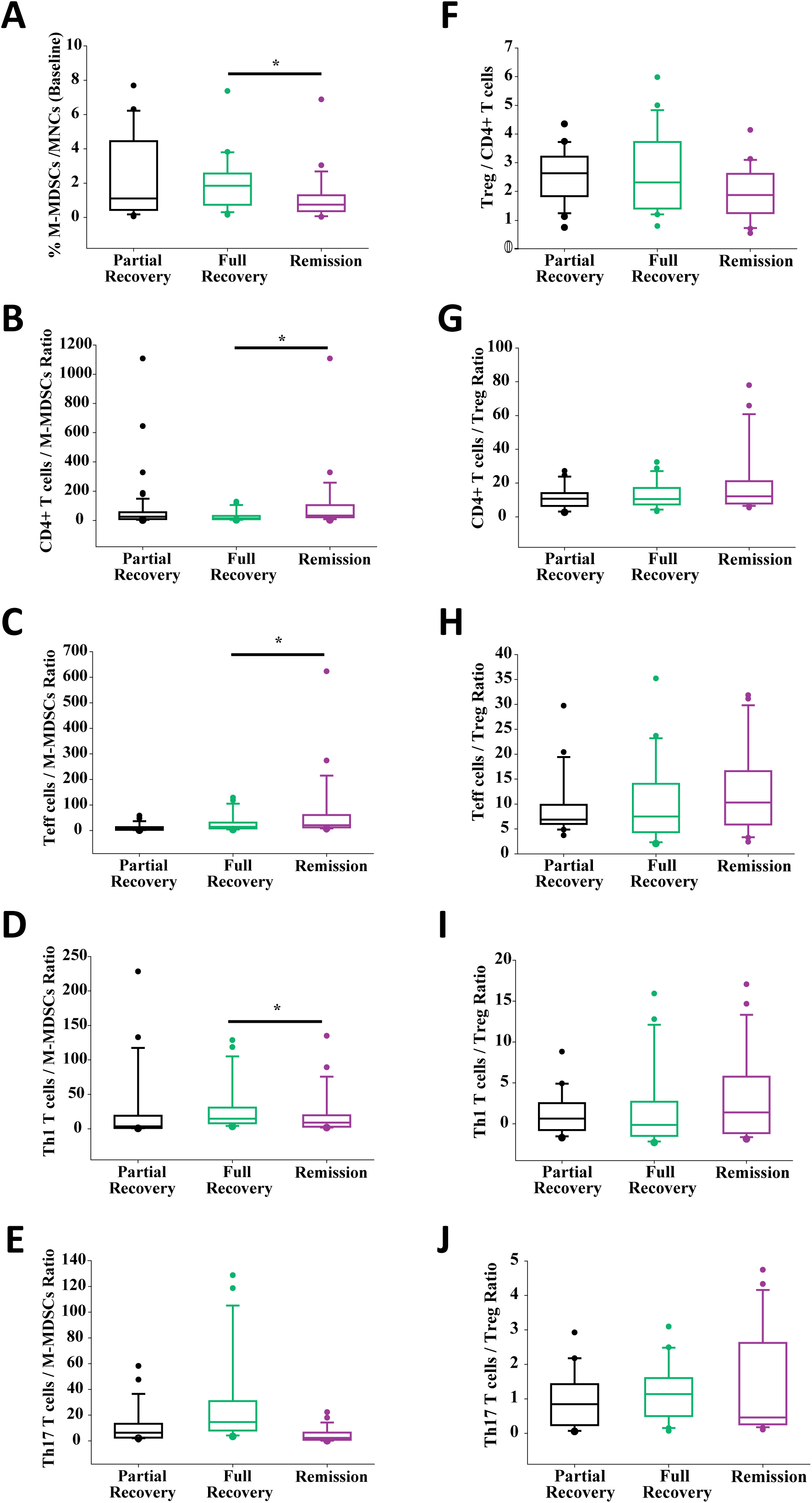
Baseline immune cell frequencies according to clinical recovery status. Patients sampled during relapse who later achieved full or partial recovery were compared with patients sampled during remission. M-MDSC levels (A), as well as ratios of CD4⁺ T cells (B), total Teff cells (C), and Th1 cells (D) to M-MDSCs, differed between patients sampled during relapse with a full recovery and those sampled during remission, whereas ratio of Th17 cells to M-MDSCs did not show significant differences (E). In contrast, neither total Treg frequencies nor effector T cell-to-Treg ratios differed among groups (F-J).

**Table 6.** Distribution of baseline cell types in relapsing MS patients with full or partial recovery and patients at remission //.

|  | **Relapse Partial**<br>**Recovery**<br>**(n = 23)** | **Relapse Full**<br>**Recovery**<br>**(n = 21)** | **Remission**<br>**(n = 24)** | **p** |
| --- | --- | --- | --- | --- |
| CD3<sup>+</sup> T cells | 58.50 (44.30-67.40) | 63.70 (48.80-68.35) | 55.10 (44.30-66.85) | 0.290 |
| CD4<sup>+</sup> T cells | 53.60 (33.10-62.70) | 47.50 (28.55-52.55) | 50.05 (45.98-54.98) | 0.278 |
| Teff | 9.65 (5.43-11.90) | 6.80 (4.68-10.79) | 9.15 (5.53-12.55) | 0.532 |
| TeffCD161<sup>+</sup> | 4.03 (2.43-5.55) | 3.35 (2.21-6.03) | 4.78 (2.54-6.58) | 0.517 |
| Th1 | 16.10 (10.60-25.80) | 16.00 (5.80-26.10) | 10.85 (5.09-22.18) | 0.368 |
| Th1.17 | 10.20 (4.81-14.50) | 6.65 (1.98-17.05) | 4.91 (2.32-9.70) | 0.204 |
| Th17 | 3.21 (1.62-6.00) | 5.51 (2.85-7.84) | 3.31 (0.91-5.01) | 0.110 |
| M-MDSCs | 1.07 (0.55-3.95) | 1.85 (0.75-2.52)* | 0.75 (0.37-1.29) | **0.042** |
| Treg | 2.63 (1.83-3.21) | 2.31 (1.43-3.64) | 1.87 (1.26-2.56) | 0.089 |
| TregCCR6<sup>+</sup> | 0.73 (0.46-1.08) | 0.69 (0.39-1.04) | 0.45 (0.22-0.79) | 0.061 |
| TregCD39<sup>+</sup> | 1.01 (0.61-1.86) | 0.89 (0.60-1.65) | 0.93 (0.45-1.57) | 0.725 |
| CD3<sup>+</sup> T cells / M-MDSCs | 45.86 (10.67-101.31) | 40.86 (21.81-72.56)* | 86.74 (44.75-165.86) | **0.032** |
| CD4<sup>+</sup> T cells / M-MDSCs | 24.06 (4.15-53.12) | 14.68 (8.41-28.75)* | 33.25 (23.08-92.03) | **0.043** |
| Teff /M-MDSCs | 15.86 (4.51-25.68) | 12.42 (6.51-22.11)* | 20.92 (12.16-56.49) | **0.032** |
| TeffCD161<sup>+</sup> / M-MDSCs | 8.19 (2.18-12.71) | 7.08 (3.36-11.39)* | 11.74 (7.16-24.06) | **0.039** |
| Th1 / M-MDSCs | 3.62 (1.24-16.78) | 3.01 (1.54-5.13)* | 9.05 (3.37-17.73) | **0.040** |
| Th1.17 / M-MDSCs | 1.75 (1.04-9.03) | 1.68 (0.92-3.64) | 4.31 (1.61-9.55) | 0.109 |
| Th17 / M-MDSCs | 0.98 (0.53-2.68) | 1.29 (0.70-2.05) | 2.34 (0.89-6.42) | 0.227 |
| CD3<sup>+</sup> T cells / Treg | 20.72 (15.51-31-77) | 30.84 (14.56-37.67) | 28.31 (16.26-39.30) | 0.386 |
| CD4<sup>+</sup> T cells / Treg | 10.77 (6.55-13.98) | 10.53 (7.37-16.96) | 12.19 (8.00-20.68) | 0.408 |
| Teff / Treg | 6.88 (6.02-9.51) | 7.49 (4.58-13.99) | 10.31 (5.97-13.83) | 0.519 |
| TeffCD161<sup>+</sup> / Treg | 3.35 (2.42-4.10) | 3.60 (2.10-6.79) | 5.08 (3.45-7.82) | 0.126 |
| Th1 / Treg | 3.08 (1.69-4.44) | 2.09 (0.93-4.40) | 3.45 (1.19-7.16) | 0.366 |
| Th1.17 / Treg | 2.27 (0.83-3.12) | 0.87 (0.44-3.50) | 1.23 (0.66-3.07) | 0.796 |
| Th17 / Treg | 0.85 (0.28-1.36) | 1.14 (0.52-1.57) | 0.50 (0.26-2.55) | 0.602 |

//The values are the median (IQR) of each group. \*p<0.05 vs. Remission

Longitudinal analysis revealed that only patients with full recovery exhibited a significant reduction in CD3^+^ T cells, Teff, TeffCD161⁺, Th17, and Th1.17 cells over the 12-month follow-up (**Figure 6A-F**). M-MDSC frequencies increased in patients sampled during relapse regardless of recovery status (**Figure 6G**), whereas Treg populations remained stable (**Figure 6H-J**). In good agreement, patients with baseline M-MDSC levels above the median showed Teff and Treg dynamics similar to those observed in patients with full recovery (**Figure 7A-I)**. Notably, no differences in demographic or baseline clinical variables were observed between both groups (**Supplementary Table 6**).

**Figure 6.**
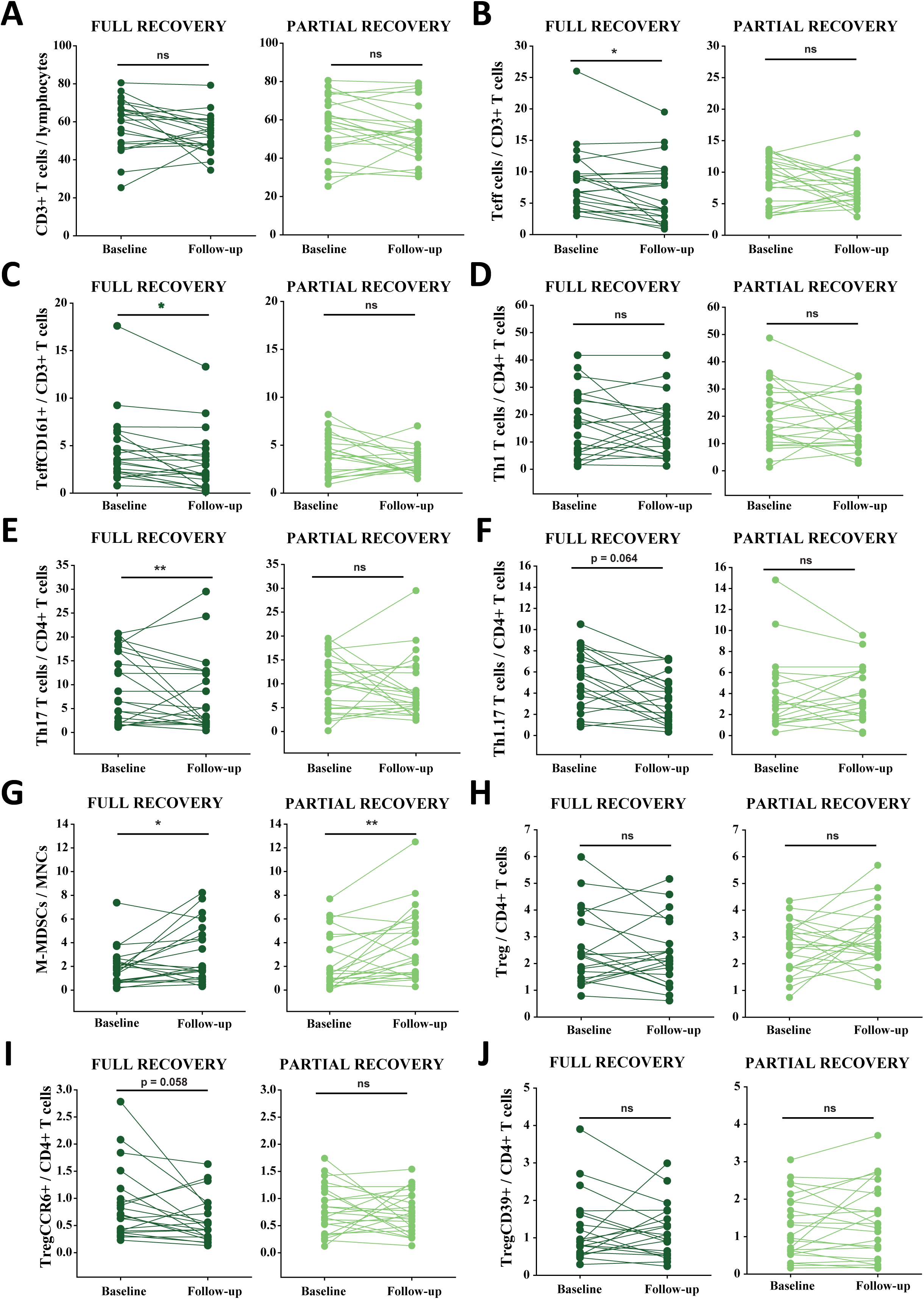
Longitudinal immune cell dynamics in patients sampled during relapse stratified by recovery outcome. Significant reductions in CD3⁺ T cells (A), total Teff (B), TeffCD161⁺ (C), Th17 (E), and Th1.17 cells (F) were observed only in patients with full recovery, whereas these changes were not detected in patients with partial recovery. Th1 cells remained stable over time (D). M-MDSC frequencies increased over time regardless of recovery (G), while total Tregs, TregCCR6⁺, and TregCD39⁺ remained stable in all patients (H–J).

**Figure 7.**
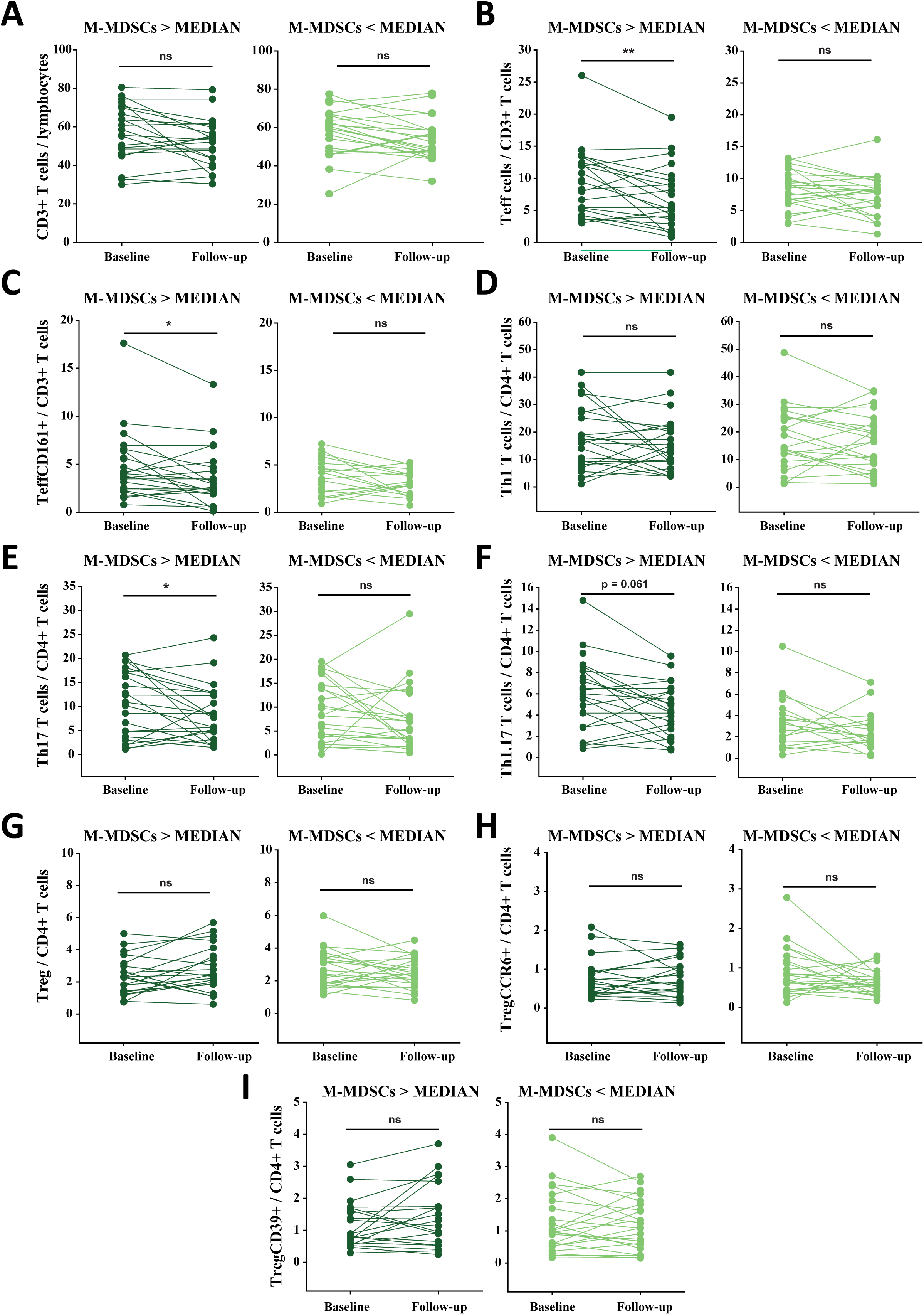
Immune cell trajectories according to baseline M-MDSC abundance in patients sampled during relapse. Patients were stratified by baseline M-MDSC levels (above *vs*. below the median) and followed for one year. Higher baseline M-MDSC levels were associated with reductions in CD3⁺ T cells, total Teff, TeffCD161⁺, and Th17 cells over time (A–C,E), whereas Th1 cells remained stable (D) and Th1.17 cells showed a trend toward reduction (F). Total Treg, TregCCR6⁺, and TregCD39⁺ frequencies remained stable (G–I). No significant longitudinal changes were observed in patients with lower baseline M-MDSC levels.

In summary, baseline M-MDSC levels, but not Treg levels, were consistently higher in RRMS patients sampled during relapse who ultimately achieved full recovery. These findings suggest that, among regulatory cell populations, M-MDSCs may serve as the most informative immunological marker for favorable relapse recovery in untreated RRMS patients.

## DISCUSSION

In this longitudinal study of untreated RRMS patients, we investigated whether peripheral immunoregulatory dynamics were associated with clinical recover following relapse by characterizing the behavior of two immune populations, M-MDSCs and Treg, over a 12-month follow-up. Our results provide insight into how M-MDSCs may shape the immunoregulatory landscape associated with relapse recovery.

One of the most salient findings was the progressive increase in circulating M-MDSCs over 12 months, irrespective of whether patients were sampled during relapse or remission. To our knowledge, this represents the first longitudinal analysis of M-MDSC levels in RRMS patients enrolled while untreated at baseline. This observation suggests that M-MDSCs may represent a sustained compensatory mechanism responding to ongoing immune activation, rather than merely acute-phase reactants. While our study focused on M-MDSCs, previous work has reported longitudinal increases in polymorphonuclear (PMN)-MDSCs in RRMS patients achieving no evidence of disease activity (NEDA-3) compared to patients with ongoing disease activity[19]. Although these findings are not directly comparable to our M-MDSC results, they support the notion that distinct MDSC subsets may expand in response to chronic immune activation and contribute to immune regulation in MS. Although some patients received corticosteroids early during follow-up, none had received corticosteroids within the 6 months preceding the 12-month follow-up, making a direct steroid-driven effect on M-MDSC levels at that time unlikely [20]. However, although most patients initiated DMT during follow-up, treatment exposure was not associated with the main longitudinal immune changes observed, and no treatment-by-time interactions were detected for any subset. These findings argue against treatment initiation as the main driver of the longitudinal increase in M-MDSCs, which is therefore more likely to reflect endogenous immune regulatory mechanisms associated with relapse evolution.

In our cohort, baseline M-MDSC levels in patients sampled during relapse were associated with subsequent immunoregulatory features, including reduced global CD4^+^ T cell frequencies, elevated Treg number, and higher FoxP3 expression at one year, suggesting a link between early M-MDSCs levels and the development of a more regulatory immune profile over time. The influence of MDSCs as promoters of Treg number and activity has been previously described in many pathological settings[18],[22], including the MS model EAE where MDSC expansion coincides with Treg increase in the spleen and/or the CNS[23],[24]. In contrast, no similar associations were observed in untreated patients sampled during remission, highlighting the importance of evaluating regulatory populations in the immediate post-relapse period.

Longitudinal analyses revealed that effector T cell dynamics were not uniform across the cohort. While several effector populations, including Teff, TeffCD161⁺, Th17 and Th1.17 cells, declined over time in patients sampled during relapse, this reduction was largely confined to patients who subsequently achieved complete clinical recovery. Previous cross-sectional studies have consistently reported increased frequencies of Th17, and Th1.17, but not Th1 subsets during MS relapses compared with remission, supporting their contribution to inflammatory disease activity [25]. However, longitudinal data capturing the evolution of these populations within the same patients, particularly in relation to clinical recovery, remain scarce. In this context, our findings extend existing knowledge by showing that the post-relapse reduction of effector T cell populations is not uniform but preferentially observed in patients achieving complete recovery. It is conceivable that these patients represent a biologically distinct subgroup with an intrinsic predisposition toward more efficient control of post-relapse inflammatory activity and favorable clinical evolution. The concomitant and sustained expansion of M-MDSCs, independent of recovery status, further suggests that peripheral immunoregulation in early RRMS is selective and context-dependent rather than reflecting a global suppression of effector responses.

Notably, in patients sampled during relapse, M-MDSC levels positively correlated with Th17 cell frequencies both at baseline and at 12-month follow-up, whereas correlations with other effector T cell subsets were weak or non-significant. Although M-MDSCs are classically regarded as suppressive cells, under certain autoimmune and inflammatory conditions they may also be associated with Th17-promoting environments [26][27]. In our cohort, the positive association observed between M-MDSCs and Th17 cells at follow-up may reflect different, non-mutually exclusive scenarios, including persistent inflammatory activity, a compensatory expansion of M-MDSCs in response to increased Th17 burden, or context-dependent functional changes in the M-MDSC compartment. These possibilities warrant further functional investigation. Finally, the longitudinal increase in Th17 cells observed in our cohort does not necessarily reflect a purely pro-inflammatory process. It may also be compatible with persistent subclinical inflammatory activity not captured by routine clinical assessment. In addition, recent evidence has highlighted the existence of IL-17-producing Treg, which display marked functional plasticity and can exert context-dependent regulatory or inflammatory roles[28]. To date, this regulatory IL-17-producing Treg subset has not been systematically explored in MS, particularly in longitudinal settings. Therefore, it is conceivable that the apparent increase in Th17 frequencies over time may partly reflect dynamic changes within the Th17 compartment itself, including shifts in the balance between pathogenic Th17 cells and IL-17-producing Treg. Such plasticity could contribute to the complex associations observed between M-MDSCs, Treg activation, and Th17 frequencies, and warrants further functional and phenotypic investigation in MS.

Regarding clinical outcomes, our data indicate that M-MDSCs, rather than Treg, were associated with full recovery after relapse. Patients who subsequently achieved complete EDSS recovery showed higher baseline M-MDSC levels compared to those sampled during remission whereas no differences were observed between full and partial recovery groups. In contrast, Treg levels and Teff/Treg ratios did not differ across recovery groups. These findings aligns with experimental models, in which M-MDSCs limit effector T cell activation and promote processes involved in the resolution of inflammatory responses[29]–[31]. Clinically, this suggests that patients with higher M-MDSC levels during relapse may possess a more effective endogenous mechanism to restrain immune hyperactivation and facilitate restoration of immune homeostasis after acute inflammatory insults. Rather than reflecting a generalized immunosuppressive state, elevated M-MDSCs may indicate a regulatory competence that supports tissue protection and functional recovery. Collectively, these observations indicate that elevated M-MDSCs may reflect a regulatory environment compatible with more efficient inflammatory control during relapse in early RRMS.

In contrast to M-MDSCs, Treg dynamics were comparatively stable over time, and baseline Treg levels did not correlate with effector T cell subsets at both baseline and 12 months later. The stability of Treg frequencies over time in our cohort differs from several previous cross-sectional studies reporting reduced Treg numbers or impaired function in MS patients[17],[22]. In an apparent contradiction, longitudinal profiling of Treg markers in MS patients from the placebo arm of a clinical trial, followed bimonthly for over a year, showed increased FoxP3 and other Treg-associated transcripts during clinical relapses compared to stable periods, suggesting dynamic modulation of the regulatory compartment over time[32]. However, our observations align with a meta-analysis that reported heterogeneous findings regarding Treg frequency and function in MS and did not identify a consistent association between Treg levels and disease activity or prognostic outcomes[33]. Collectively, these data underscore the variability and context- dependence of Treg dynamics in MS and suggest that peripheral Treg frequency may not serve as a reliable longitudinal biomarker, unlike M-MDSCs.

Several limitations of this study should be acknowledged. First, the cohort size, while well-characterized, is relatively small, which may limit generalizability and the detection of subtle immunological differences. Second, functional assays directly assessing suppressive activity of M-MDSCs and Treg were not carried out, limiting mechanistic interpretation. Third, our follow-up was restricted to one year; longer longitudinal studies are required to assess the persistence of these regulatory dynamics and their relationship with long-term disability and therapeutic responses. Finally, although all patients were untreated at baseline and many initiated DMT during follow-up, no treatment-by-time interactions were detected, arguing against treatment exposure as the main driver of the longitudinal immune changes observed.

In conclusion, our findings support the existence of a distinct immunoregulatory profile associated with favorable relapse recovery in early RRMS. Higher baseline M-MDSC levels were linked to subsequent changes in the regulatory immune compartment and to complete clinical recovery after relapse, whereas Treg dynamics remained largely stable and were not associated with recovery outcomes. These observations suggest that M-MDSCs may reflect an intrinsic regulatory competence capable of restraining post-relapse immune hyperactivation. Further studies will be required to determine the functional mechanisms underlying these associations and their potential implications for understanding immune regulation during relapse recovery.

## Data Availability

All data produced in the present study are available upon reasonable request to the authors

## Figure legends

**Supplementary Figure 1. Flow cytometry gating strategy for the identification of M-MDSCs.** After exclusion of doublets and dead cells based on Zombie NIR staining, live mononuclear cells (MNCs) were gated. Immature myeloid cells were then identified within the monocyte population as CD33⁺HLA-DR⁻^/low^ cells. Finally, CD14 and CD15 expression was assessed within the CD33⁺HLA-DR⁻^/low^ population to define M-MDSCs as CD14⁺CD15⁻ cells.

**Supplementary Figure 2. Flow cytometry gating strategy for effector T cell subsets.** After exclusion of doublets and dead cells based on Zombie NIR staining, viable cells were selected according to forward scatter (FSC) and side scatter (SSC) characteristics. T cells were identified as CD3⁺ cells, and T helper cells were defined as CD4⁺ cells. CD4⁺ T cell subsets were categorized as effector T cells (Teff; CCR2⁺CCR5⁺), TeffCD161⁺ cells (CCR2⁺CCR5⁺CD161⁺), Th1 cells (CXCR3⁺CCR6⁻), Th1.17 cells (CXCR3⁺CCR6⁺), and Th17 cells (CXCR3⁻CCR6⁺CD161⁺).

**Supplementary Figure 3. Flow cytometry gating strategy for the analysis of Treg.** After exclusion of doublets and dead cells based on Zombie NIR staining, viable cells were selected according to forward scatter (FSC) and side scatter (SSC) characteristics. T cells were identified as CD3⁺ cells, and T helper cells were defined as CD4⁺ cells. CD4⁺ T cells were then categorized as total Treg (CD25^hi^CD127^low^FoxP3⁺), Treg with CNS migratory potential (CD25^hi^CD127^low^FoxP3⁺CCR6⁺), and Treg with high immunosuppressive activity (CD25^hi^CD127^low^FoxP3⁺CD39⁺).

## Acknowledgments

The authors would like to express our gratitude to all MS patient volunteers from the three participant hospitals, and to Ángela Marquina Rodríguez and María José González López at the Flow Cytometry Core Facility of the *Hospital Nacional de Paraplejicos* for their technical assistance.

## Ethical Considerations

The Clinical Investigation Ethical Committee review board of the *Complejo Hospitalario Universitario de Toledo approved* the study (No 349).

## Consent to Participate

All participants provided informed written consent.

## Consent for publication

Not applicable.

## Data availability statement

Anonymized data will be shared upon reasonable request from any qualified investigator, after approval of the ethical committee of the *Complejo Hospitalario Universitario de Toledo*.

## References

1. Thompson AJ, Baranzini SE, Geurts J, Hemmer B, Ciccarelli O. Multiple sclerosis. Lancet. 2018; 391(10130):1622–1636. Available at: https://linkinghub.elsevier.com/retrieve/pii/S0140673618304811.

2. Lublin FD, Häring DA, Ganjgahi H, et al. How patients with multiple sclerosis acquire disability. Brain. 2022; 145(9):3147–3161. Available at: https://academic.oup.com/brain/article/145/9/3147/6519354.

3. Dzau W, Sharmin S, Patti F, et al. Risk of secondary progressive multiple sclerosis after early worsening of disability. J. Neurol. Neurosurg. Psychiatry. 2023; 94(12):984–991. Available at: https://jnnp.bmj.com/lookup/doi/10.1136/jnnp-2023-331748.

4. Lublin FD, Baier M, Cutter G. Effect of relapses on development of residual deficit in multiple sclerosis. Neurology. 2003; 61(11):1528–1532. Available at: https://www.neurology.org/doi/10.1212/01.WNL.0000096175.39831.21.

5. Nemecek A, Zimmermann H, Rübenthaler J, et al. Flow cytometric analysis of T cell/monocyte ratio in clinically isolated syndrome identifies patients at risk of rapid disease progression. Mult. Scler. J. 2016; 22(4):483–493. Available at: https://journals.sagepub.com/doi/10.1177/1352458515593821.

6. Rostami A, Ciric B. Role of Th17 cells in the pathogenesis of CNS inflammatory demyelination. J. Neurol. Sci. 2013; 333(1–2):76–87. Available at: https://linkinghub.elsevier.com/retrieve/pii/S0022510X13001081.

7. van Langelaar J, van der Vuurst de Vries RM, Janssen M, et al. T helper 17.1 cells associate with multiple sclerosis disease activity: perspectives for early intervention. Brain. 2018; 141(5):1334–1349. Available at: https://academic.oup.com/brain/article/141/5/1334/4961484.

8. Kitz A, Singer E, Hafler D. Regulatory T Cells: From Discovery to Autoimmunity. Cold Spring Harb. Perspect. Med. 2018; 8(12):a029041. Available at: http://perspectivesinmedicine.cshlp.org/lookup/doi/10.1101/cshperspect.a029041.

9. Jiang Q, Duan J, Kaer L Van, Yang G. The Role of Myeloid-Derived Suppressor Cells in Multiple Sclerosis and Its Animal Model. Aging Dis. 2023. Available at: http://www.aginganddisease.org/EN/10.14336/AD.2023.0323-1.

10. Radojević D, Bekić M, Gruden-Movsesijan A, et al. Myeloid-derived suppressor cells prevent disruption of the gut barrier, preserve microbiota composition, and potentiate immunoregulatory pathways in a rat model of experimental autoimmune encephalomyelitis. Gut Microbes. 2022; 14(1). Available at: https://www.tandfonline.com/doi/full/10.1080/19490976.2022.2127455.

11. Ortega MC, Lebrón-Galán R, Machín-Díaz I, et al. Central and peripheral myeloid-derived suppressor cell-like cells are closely related to the clinical severity of multiple sclerosis. Acta Neuropathol. 2023; (0123456789). Available at: 10.1007/s00401-023-02593-x.

12. Thompson AJ, Banwell BL, Barkhof F, et al. Diagnosis of multiple sclerosis: 2017 revisions of the McDonald criteria. Lancet Neurol. 2018; 17(2):162–173. Available at: https://linkinghub.elsevier.com/retrieve/pii/S1474442217304702.

13. Ramo-Tello C, Blanco Y, Brieva L, et al. Recommendations for the Diagnosis and Treatment of Multiple Sclerosis Relapses. J. Pers. Med. 2021; 12(1):6. Available at: https://www.mdpi.com/2075-4426/12/1/6.

14. Gu J, Ni X, Pan X, et al. Human CD39hi regulatory T cells present stronger stability and function under inflammatory conditions. Cell. Mol. Immunol. 2017; 14(6):521–528. Available at: https://www.nature.com/articles/cmi201630.

15. Verreycken J, Baeten P, Broux B. Regulatory T cell therapy for multiple sclerosis: Breaching (blood-brain) barriers. Hum. Vaccin. Immunother. 2022; 18(7). Available at: https://www.tandfonline.com/doi/full/10.1080/21645515.2022.2153534.

16. Sato W, Tomita A, Ichikawa D, et al. CCR2+CCR5+ T Cells Produce Matrix Metalloproteinase-9 and Osteopontin in the Pathogenesis of Multiple Sclerosis. J. Immunol. 2012; 189(10):5057–5065. Available at: https://academic.oup.com/jimmunol/article/189/10/5057/7997831.

17. Mexhitaj I, Nyirenda MH, Li R, et al. Abnormal effector and regulatory T cell subsets in paediatric-onset multiple sclerosis. Brain. 2019; 142(3):617–632. Available at: https://academic.oup.com/brain/article/142/3/617/5316384.

18. Tamberi L, Belloni A, Pugnaloni A, et al. The Influence of Myeloid-Derived Suppressor Cell Expansion in Neuroinflammation and Neurodegenerative Diseases. Cells. 2024; 13(7):643. Available at: https://www.mdpi.com/2073-4409/13/7/643.

19. Knier B, Hiltensperger M, Sie C, et al. Myeloid-derived suppressor cells control B cell accumulation in the central nervous system during autoimmunity. Nat. Immunol. 2018; 19(12):1341–1351. Available at: https://www.nature.com/articles/s41590-018-0237-5.

20. Wang Z, Zheng G, Li G, et al. Methylprednisolone alleviates multiple sclerosis by expanding myeloid-derived suppressor cells via glucocorticoid receptor β and S100A8/9 up-regulation. J. Cell. Mol. Med. 2020; 24(23):13703–13714. Available at: https://onlinelibrary.wiley.com/doi/10.1111/jcmm.15928.

21. Cantoni C, Cignarella F, Ghezzi L, et al. Mir-223 regulates the number and function of myeloid-derived suppressor cells in multiple sclerosis and experimental autoimmune encephalomyelitis. Acta Neuropathol. 2017; 133(1):61–77. Available at: http://link.springer.com/10.1007/s00401-016-1621-6.

22. Calahorra L, Camacho-Toledano C, Serrano-Regal MP, Ortega MC, Clemente D. Regulatory Cells in Multiple Sclerosis: From Blood to Brain. Biomedicines. 2022; 10(2):335. Available at: https://www.mdpi.com/2227-9059/10/2/335.

23. Tanwar S, Oguz C, Metidji A, et al. Type I IFN signaling in T regulatory cells modulates chemokine production and myeloid derived suppressor cells trafficking during EAE. J. Autoimmun. 2020; 115:102525. Available at: https://linkinghub.elsevier.com/retrieve/pii/S0896841120301499.

24. Lamba T, Zafar MA, Shaaz M, et al. Mycobacterium tuberculosis Acr1 Protein Mitigates Experimental Autoimmune Encephalomyelitis Symptoms by Generating Myeloid-Derived Suppressor Cells and Regulatory T Cells. Immunology. 2026; 177(2):370–383. Available at: https://onlinelibrary.wiley.com/doi/10.1111/imm.70050.

25. Kalra S, Lowndes C, Durant L, et al. Th17 cells increase in RRMS as well as in SPMS, whereas various other phenotypes of Th17 increase in RRMS only. Mult. Scler. J. - Exp. Transl. Clin. 2020; 6(1). Available at: https://journals.sagepub.com/doi/10.1177/2055217319899695.

26. Glenn JD, Liu C, Whartenby KA. Frontline Science: Induction of experimental autoimmune encephalomyelitis mobilizes Th17-promoting myeloid derived suppressor cells to the lung. J. Leukoc. Biol. 2019; 105(5):829–841. Available at: https://academic.oup.com/jleukbio/article/105/5/829/6935635.

27. Sanchez-Pino MD, Dean MJ, Ochoa AC. Myeloid-derived suppressor cells (MDSC): When good intentions go awry. Cell. Immunol. 2021; 362:104302. Available at: https://linkinghub.elsevier.com/retrieve/pii/S0008874921000216.

28. Cui H, Wang N, Li H, et al. The dynamic shifts of IL-10-producing Th17 and IL-17-producing Treg in health and disease: a crosstalk between ancient ‘Yin-Yang’ theory and modern immunology. Cell Commun. Signal. 2024; 22(1):99. Available at: https://biosignaling.biomedcentral.com/articles/10.1186/s12964-024-01505-0.

29. Alabanza LM, Esmon NL, Esmon CT, Bynoe MS. Inhibition of Endogenous Activated Protein C Attenuates Experimental Autoimmune Encephalomyelitis by Inducing Myeloid-Derived Suppressor Cells. J. Immunol. 2013; 191(7):3764–3777. Available at: https://academic.oup.com/jimmunol/article/191/7/3764/7989681.

30. Moliné-Velázquez V, Ortega MC, Vila del Sol V, et al. The synthetic retinoid Am80 delays recovery in a model of multiple sclerosis by modulating myeloid-derived suppressor cell fate and viability. Neurobiol. Dis. 2014; 67:149–164. Available at: https://linkinghub.elsevier.com/retrieve/pii/S0969996114000783.

31. Melero-Jerez C, Suardíaz M, Lebrón-Galán R, et al. The presence and suppressive activity of myeloid-derived suppressor cells are potentiated after interferon-β treatment in a murine model of multiple sclerosis. Neurobiol. Dis. 2019; 127:13–31. Available at: https://linkinghub.elsevier.com/retrieve/pii/S0969996119300439.

32. Dalla Libera D, Di Mitri D, Bergami A, et al. T Regulatory Cells Are Markers of Disease Activity in Multiple Sclerosis Patients Jacobson S, ed. PLoS One. 2011; 6(6):e21386. Available at: https://dx.plos.org/10.1371/journal.pone.0021386.

33. Noori-Zadeh A, Mesbah-Namin SA, Bistoon-beigloo S, et al. Regulatory T cell number in multiple sclerosis patients: A meta-analysis. Mult. Scler. Relat. Disord. 2016; 5:73–76. Available at: https://linkinghub.elsevier.com/retrieve/pii/S2211034815300213.

