## Supplementary Information for "Monocytic myeloid-derived suppressor cells, but not regulatory T cells, track immunoregulatory dynamics and relapse recovery in early RRMS"

**Supplementary Table 1. Antibody panels used in BD Canto II Cytometer***

| Marker | Filter | Effector T cell panel | Clon | Regulatory T cell panel | Clon | MDSC panel | Clon |
|---|---|---|---|---|---|---|---|
| **Blue(488nm)** | | | | | | | |
| FITC | 530/30 | CXCR3 | G025H7 | CD127 | hIL-7R-M21 | CD15 | HI98 |
| PE | 585/42 | CCR6 | G034E3 | CCR6 | G034E3 | Lox-1 | 15C4 |
| Per-CP Cy5.5 | 670 LP | CD161 | HP-3G10 | CD25 | M-A251 | CD14 | MφP9 |
| PE-Cy7 | 780/60 | CD3 | UCT1 | CD3 | UCT1 | CD11b | ICRF44 |
| **Red (633 nm)** | | | | | | | |
| APC | 660/20 | CCR5 | 2D/CCR5 | _ | _ | CD33 | WM53 |
| Alexa Fluor 647 | 660/20 | _ | _ | FoxP3 | 236A/E7 | _ | _ |
| NIR | 780/60 | Zombie | | Zombie | | Zombie | |
| **Violet (405 nm)** | | | | | | | |
| PB | 450/50 | CD4 | OKT4 | CD4 | OKT4 | _ | _ |
| BV421 | 450/50 | _ | _ | _ | _ | HLA-DR | G46-6 |
| BV510 | 530/30 | CCR2 | K036C2 | CD39 | A1 | _ | _ |

**CD11b, CD14, CD15, CD33, HLA-DR, CD3, CD127, CD25, FoxP3, CCR5 were from BD Pharmigen. CD4, CCR6, CD39, CD161, and CCR2 were from Biolegend.*

**Supplementary Table 2. Phenotype of the analyzed leukocyte subsets**

| Leukocyte subpopulation | Phenotype |
|---|---|
| **Effector T cells** | |
| $CD3^+$ T cells | $CD3^+$ |
| $CD4^+$ T cells | $CD3^+CD4^+$ |
| Th1 | $CD3^+$ $CD4^+CCR6^-$ $CXCR3^+$ |
| Th17 | $CD3^+$ $CD4^+CCR6^+$ $CXCR3^-$ $CD161^+$ |
| Th1.17 | $CD3^+$ $CD4^+CCR6^+$ $CXCR3^+$ |
| Effector T cells (Teff) | $CD3^+$ $CD4^+CCR2^+$ $CCR5^+$ |
| Teff $CD161^+$ T cells | $CD3^+$ $CD4^+CCR2^+$ $CCR5^+$ $CD161^+$ |
| **Regulatory Tcells (Treg)** | |
| Treg | $CD3^+$ $CD4^+$ $CD127^{lo}$ $CD25^{hi}$ $FoxP3^+$ |
| Treg $CD39^+$ | $CD3^+$ $CD4^+$ $CD127^{lo}$ $CD25^{hi}$ $FoxP3^+$ $CD39^+$ |
| Treg $CCR6^+$ | $CD3^+$ $CD4^+$ $CD127^{lo}$ $CD25^{hi}$ $FoxP3^+$ $CCR6^+$ |
| **Myeloid-derived suppressor cells (MDSCs)** | |
| Monocytic- MDSCs (M-MDSCs) | $CD11b^+$ $CD33^+$ HLA-$DR^{-/low}$ $CD14^+$ $CD15^-$ |

**Supplementary Table 3. Distribution of cell frequencies at one-year follow-up of treated/untreated MS patients sampled during relapse**

| | | **Relapse treated (n = 34)** | **Relapse untreated (n = 11)** | ***p*** |
|---|---|---|---|---|
| **ONE YEAR FOLLOW-UP** | M-MDSCs/MNCs | 2.55 (0.97-5.35) | 1.82 (1.60-2.67) | 0.704 |
| | $CD3^{+}$T cells | 52.80 (44.20-58.30) | 54.70 (43.88-58.60) | 0.917 |
| | $CD4^{+}$T cells | 40.60 (31.40-55.60) | 39.20 (37.53-52.65) | 0.781 |
| | Teff | 5.79 (4.01-8.14) | 8.97 (6.97-10.28) | **0.031** |
| | $TeffCD161^{+}$ | 2.62 (1.79-3.55) | 4.00 (2.59-4.63) | 0.174 |
| | Th1 | 14.15 (9.23-20.20) | 16.60 (6.27-23.75) | 0.775 |
| | Th1.17 | 5.59 (3.16-12.30) | 6.93 (2.92-11.63) | 0.905 |
| | Th17 | 3.13 (1.75-4.80) | 2.93 (1.92-5.59) | 0.884 |
| | Treg | 2.50 (2.01-3.60) | 1.85 (1.23-2.58) | **0.018** |
| | $TregCCR6^{+}$ | 0.62 (0.42-0.92) | 0.58 (0.37-0.88) | 0.702 |
| | $TregCD39^{+}$ | 1.30 (0.55-1.91) | 0.89 (0.63-1.55) | 0.331 |

*[¥]The values are the median (IQR) of each group.*

**Supplementary Table 4. Distribution of effector T cells/TregCCR6$^+$ ratios in MS patients**//

| | | Relapse (n = 45) | Remission (n = 24) | p |
|---|---|---|---|---|
| **BASELINE** | CD3$^+$T cells / Treg | 81.31 (47.68-156.51) | 109.45 (63.65-350.00) | 0.063 |
| | CD4$^+$ T cells / Treg | 63.06 (40.67-87.96) | 101.80 (67.47-242.59) | **<0.001** |
| | Teff / Treg | 29.60 (14.23-51.63) | 43.79 (24.52-59.12) | 0.059 |
| | TeffCD161$^+$ / Treg | 13.05 (7.60-23.05) | 22.34 (11.39-34.00) | **0.018** |
| | Th1 / Treg | 18.41 (11.30-41.54) | 26.39 (12.56-63.11) | 0.217 |
| | Th1.17 / Treg | 10.02 (6.39-19.17) | 11.73 (8.61-18.65) | 0.517 |
| | Th17 / Treg | 5.75 (2.65-11.19) | 6.03 (3.93-11.08) | 0.846 |
| **ONE YEAR FOLLOW-UP** | CD3$^+$ T cells / Treg | 98.10 (50.39-129.42) | 84.97 (52.89-166.79) | 0.674 |
| | CD4$^+$ T cells / Treg | 69.41 (47.41-102.25) | 104.87 (52.90-179.17) | 0.060 |
| | Teff / Treg | 23.18 (15.35-47.25) | 30.37 (19.39-52.27) | 0.404 |
| | TeffCD161$^+$ / Treg | 10.35 (6.78-19.31) | 13.37 (8.08-22.24) | 0.267 |
| | Th1 / Treg | 19.62 (12.62-46.87) | 35.36 (13.67-55.00) | 0.259 |
| | Th1.17 / Treg | 9.77 (5.14-18.11) | 9.11 (3.72-17.71) | 0.467 |
| | Th17 / Treg | 4.65 (3.04-10.57) | 4.71 (2.65-9.58) | 0.741 |

*//The values are the median (IQR) of each group.*

**Supplementary Table 5. Distribution of effector T cell/TregCD39$^+$ ratios in MS patients**[//]

| | | Relapse (n = 45) | Remission (n = 24) | p |
|---|---|---|---|---|
| **BASELINE** | CD3$^+$ T cells / Treg | 63.77 (32.91-95.06) | 54.75 (33.26-110.66) | 0.899 |
| | CD4$^+$ T cells / Treg | 45.18 (21.28-77.21) | 62.02 (27.85-117.27) | 0.202 |
| | Teff / Treg | 21.27 (10.19-35.81) | 19.35 (11.29-43.00) | 0.846 |
| | TeffCD161$^+$ / Treg | 7.62 (4.71-15.82) | 9.04 (5.74-20.65) | 0.544 |
| | Th1 / Treg | 12.62 (7.01-27.21) | 14.16 (9.45-22.91) | 0.899 |
| | Th1.17 / Treg | 7.28 (3.56-16.40) | 5.09 (3.40-9.37) | 0.295 |
| | Th17 / Treg | 4.16 (1.93-9.20) | 2.19 (1.05-7.71) | 0.230 |
| **ONE YEAR FOLLOW-UP** | CD3$^+$T cells / Treg | 43.49 (27.29-88.55) | 35.97 (25.18-68.07) | 0.674 |
| | CD4$^+$ T cells / Treg | 43.61 (47.41-102.25) | 40.57 (25.48-109.39) | 0.640 |
| | Teff / Treg | 12.10 (7.87-31.49) | 12.36 (8.65-20.86) | 0.718 |
| | Teff CD161$^+$/ Treg | 4.23 (3.29-13.38)* | 5.25 (4.20-10.72) | 0.350 |
| | Th1 / Treg | 12.55 (6.06-24.69) | 13.76 (5.98-33.45) | 0.664 |
| | Th1.17 / Treg | 5.12 (2.72-11.65) | 3.60 (1.69-8.42) | 0.259 |
| | Th17 / Treg | 2.68 (1.43-5.70) | 1.73 (1.18-4.78) | 0.458 |

[//]*The values are the median (IQR) of each group. *p<0.05 vs. baseline*

**Supplementary Table 6. Clinical characteristics of relapsing MS patients according to the level of M-MDSCs at baseline**

| | **M-MDSCs > MEDIAN (n = 22)** | **M-MDSCs < MEDIAN (n = 23)** |
|---|---|---|
| Female (%)$^{-}$ | 14 (63.64) | 14 (66.87) |
| Age (year)$^{¥}$ | 37 (30-44) | 35 (29-42) |
| Elapsed days from relapse | 10.68 ± 1.09 | 11.74 ± 1.38 |
| Baseline EDSS$^{//}$ | 3 (2-3) | 3 (2-4) |
| 1 year follow-up EDSS$^{//}$ | 0 (0-1) | 1 (0-1.5) |
| Baseline No. of brain MRI $Gd^{+}$ lesions$^{¥}$ | 2.36 ± 0.58 | 2.09 ± 0.58 |
| 1 year No. of brain MRI $Gd^{+}$ lesions$^{¥}$ | 0.32 ± 0.19 | 0.34 ± 0.07 |
| Baseline No. of brain MRI lesions on T2$^{\&¥}$ | 26.65 ± 4.96 | 24.39 ± 4.13 |
| 0-9 lesions | 5 (22.73) | 5 (21.74) |
| ≥ 10 lesions | 17 (77.27) | 18 (78.26) |
| DMTs used during follow-up (%) | | |
| No treatment | 4 (18.18) | 7 (30.43) |
| Injectable DMTs[1] | 4 (18.18) | 4 (17.39) |
| Oral DMTs[2] | 11 (50.00) | 11 (47.83) |
| Monoclonal antibodies[3] | 3 (13.64) | 1 (4.35) |

*¥The values are the mean ± SEM of each group.*

*//The values are the median (IQR).*

*⁻The value is the percentage of women in each group, and a $Xi^2$ exact test was used to compare two proportions.*

*&The value is the number (percentage) of patients in each group, and a Fisher–Freeman–Halton exact test was used to compare the proportions.[1] Injectable DMTs: glatiramer acetate, all interferon-β formulations.*

*[2] Oral DMTs: fumarates, teriflunomide, fingolimod, cladribine.*

*[3] Monoclonal antibodies: natalizumab, alemtuzumab.*

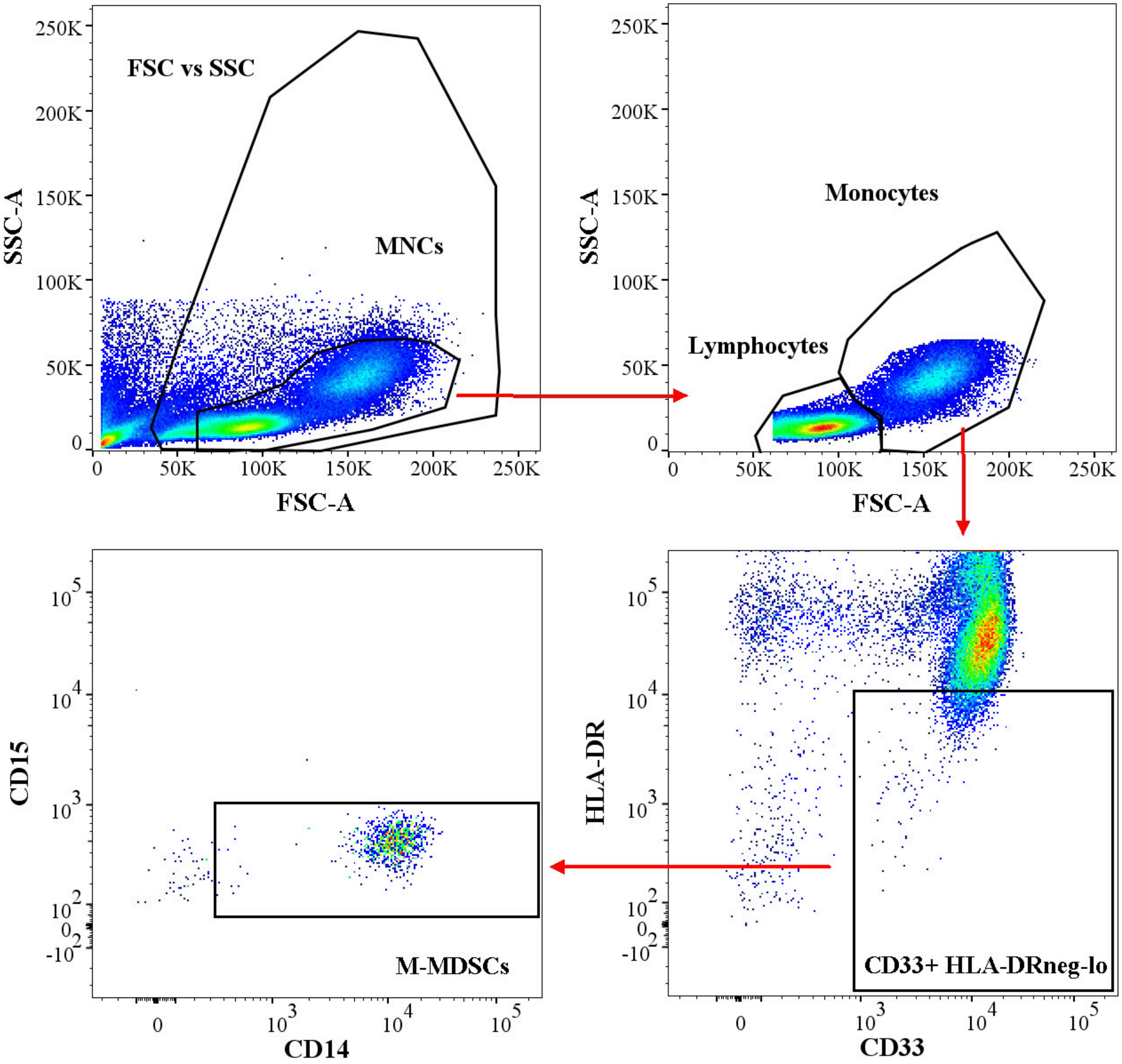

FSC vs SSC
MNCs
SSC-A
FSC-A
Monocytes
Lymphocytes
CD15
CD14
M-MDSCs
HLA-DR
CD33
CD33+ HLA-DRneg-lo

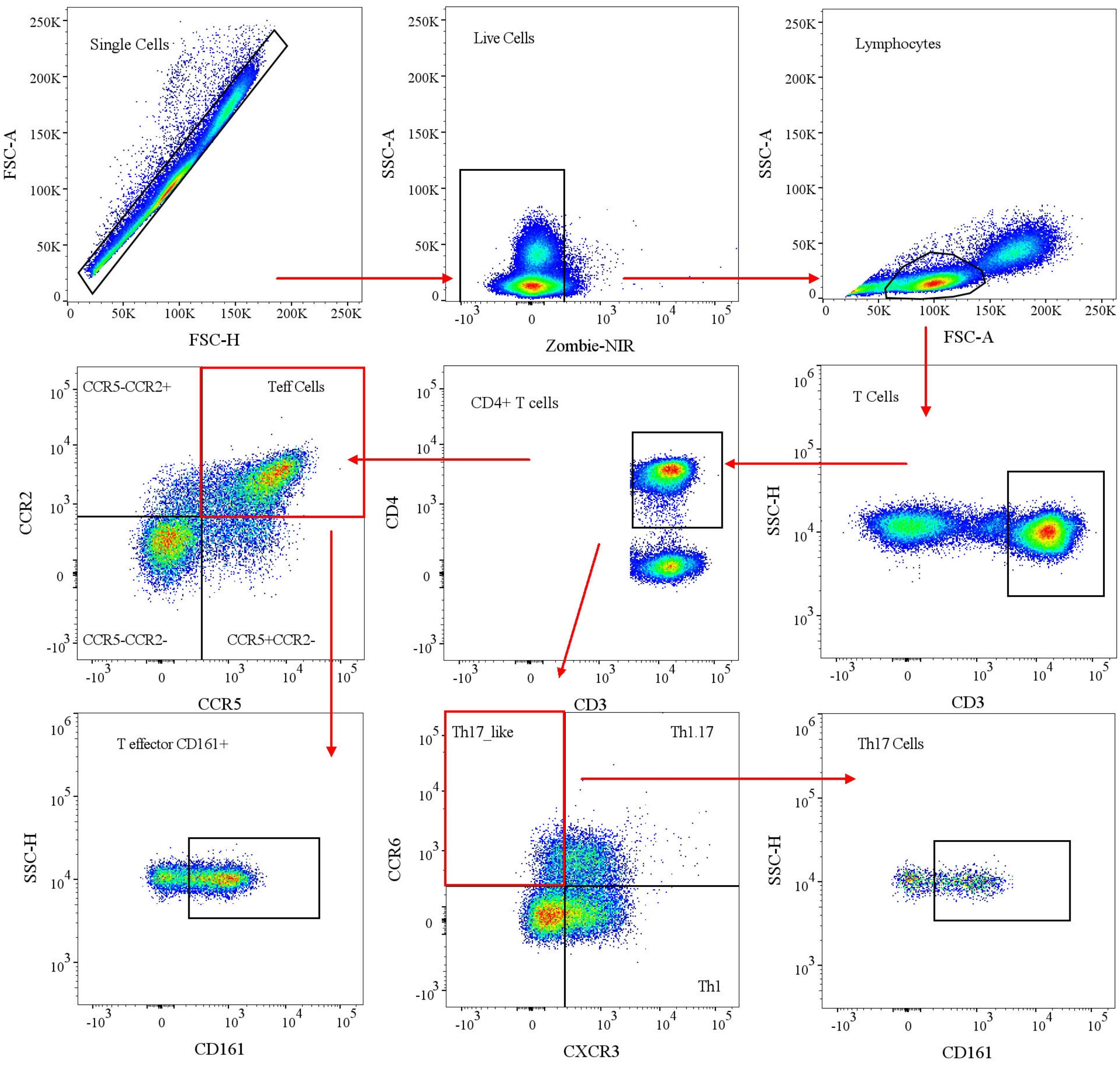

Single Cells
FSC-A
FSC-H
Live Cells
SSC-A
Zombie-NIR
Lymphocytes
SSC-A
FSC-A
CCR5-CCR2+
Teff Cells
CCR5-CCR2-
CCR5+CCR2-
CCR2
CCR5
CD4+ T cells
CD4
CD3
T Cells
SSC-H
CD3
T effector CD161+
SSC-H
CD161
Th17_like
Th1.17
Th1
CCR6
CXCR3
Th17 Cells
SSC-H
CD161

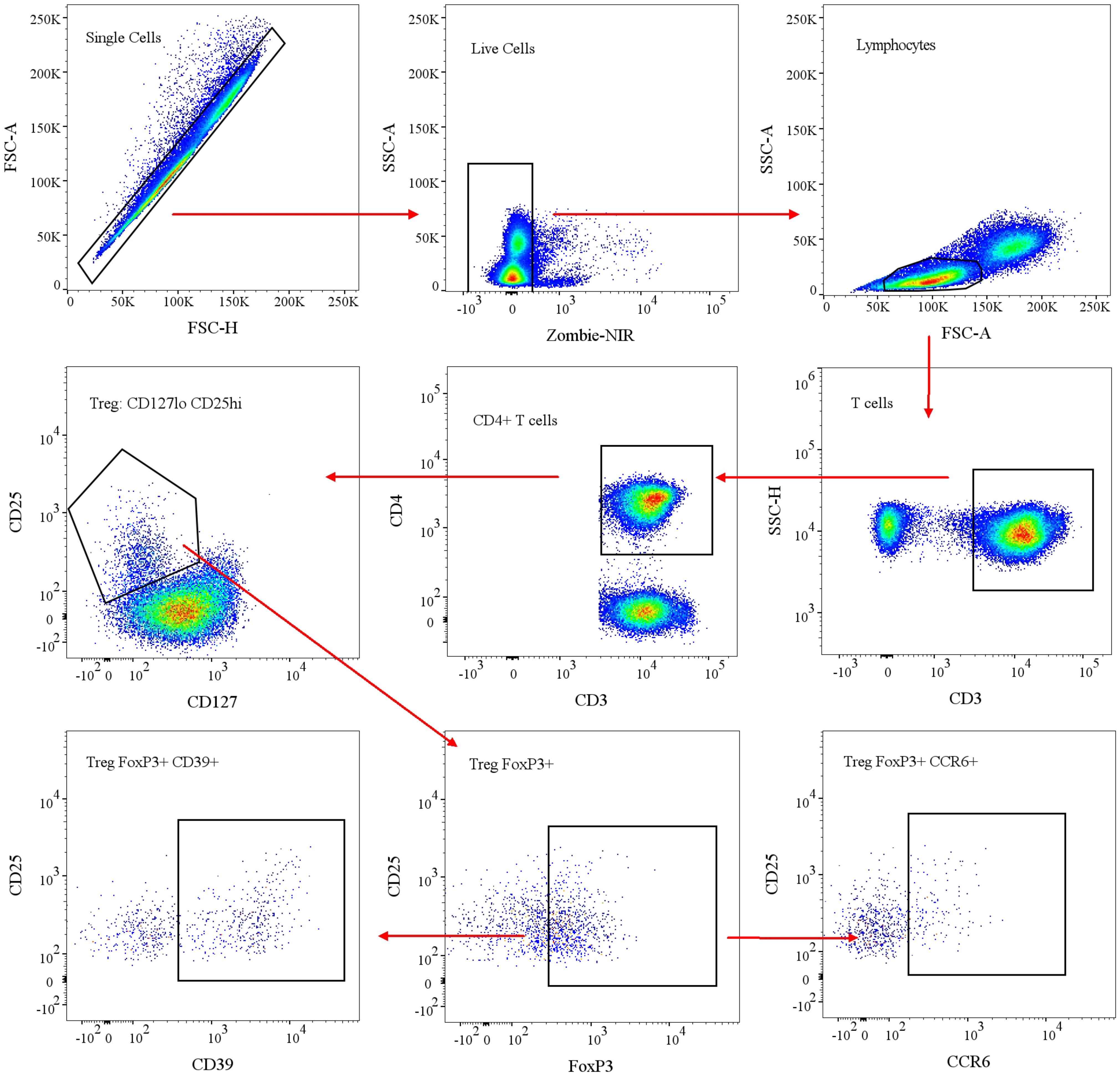
Single Cells
FSC-A
FSC-H
Live Cells
SSC-A
Zombie-NIR
Lymphocytes
SSC-A
FSC-A
Treg: CD127lo CD25hi
CD25
CD127
CD4+ T cells
CD4
CD3
T cells
SSC-H
CD3
Treg FoxP3+ CD39+
CD25
CD39
Treg FoxP3+
CD25
FoxP3
Treg FoxP3+ CCR6+
CD25
CCR6
